# Quantifying the impact of pre-vaccination titre and vaccination history on influenza vaccine immunogenicity

**DOI:** 10.1101/2024.01.24.24301614

**Authors:** David Hodgson, Stephany Sánchez-Ovando, Louise Carolan, Yi Liu, A. Jessica Hadiprodjo, Annette Fox, Sheena G. Sullivan, Adam J. Kucharski

## Abstract

Epidemiological studies suggest that heterogeneity in influenza vaccine antibody response is associated with host factors, including pre-vaccination immune status, age, gender, and vaccination history. However, the pattern of reported associations varies between studies. To better understand the underlying influences on antibody responses, we combined host factors and vaccine-induced in-host antibody kinetics from a cohort study conducted across multiple seasons with a unified analysis framework. We developed a flexible individual-level Bayesian model to estimate associations and interactions between host factors, including pre-vaccine HAI titre, age, sex, vaccination history and study setting, and vaccine-induced HAI titre antibody boosting and waning. We applied the model to derive population-level and individual effects of post-vaccine antibody kinetics for vaccinating and circulating strains for A(H1N1) and A(H3N2) influenza subtypes. We found that post-vaccine HAI titre dynamics were significantly influenced by pre-vaccination HAI titre and vaccination history and that lower pre-vaccination HAI titre results in longer durations of seroprotection (HAI titre equal to 1:40 or higher). Consequently, for A(H1N1), our inference finds that the expected duration of seroprotection post-vaccination was 171 (95% Posterior Predictive Interval[PPI] 128–220) and 159 (95% PPI 120–200) days longer for those who are infrequently vaccinated (<2 vaccines in last five years) compared to those who are frequently vaccinated (2 or more vaccines in the last five years) at pre-vaccination HAI titre values of 1:10 and 1:20 respectively. In addition, we found significant differences in the empirical distributions that describe the individual-level duration of seroprotection for A(H1N1) circulating strains. In future, studies that rely on serological endpoints should include the impact of pre-vaccine HAI titre and prior vaccination status on seropositivity and seroconversion estimates, as these significantly influence an individual’s post-vaccination antibody kinetics.

## INTRODUCTION

The global burden of influenza-associated acute respiratory disease is substantial, annually causing 5–6 million hospitalised respiratory cases and 290,000–650,000 deaths.[1,2] Vaccination remains the primary method to control seasonal influenza disease and transmission. The WHO-recommends trivalent influenza vaccines containing candidate vaccine viruses representing recent A(H1N1)pdm09, A(H3N2), and B/Victoria viruses. Recommendations for quadrivalent vaccines have previously also included a B/Yamagata virus. These recommendations are made each year roughly 6 months prior to national influenza vaccination campaigns. Vaccination campaigns typically target individuals at the highest risk of disease complications and those who are the main drivers of transmission within the population.[3] Although vaccination is a valuable preventative tool, the propensity for circulating influenza viruses to undergo antigenic evolution and amino acid substitutions that can rise in antigenic sites when viruses are propagated in eggs, means that vaccine-induced antibodies from egg-grown viruses may recognise circulating viruses poorly, resulting in low vaccine effectiveness.[4–7] Host factors also impact vaccine effectiveness, including genetic and epigenetic factors and underlying immune profile at vaccination.[8–15] Quantifying how these host factors interact to influence vaccine effectiveness is important for understanding which groups will benefit most from vaccination and help further understand the biological mechanisms that cause heterogeneity in protection from disease.

Cohort or case-control studies can measure the effectiveness of influenza vaccination by relying on case-incidence rates. However, these studies can miss non-healthcare-seeking infections and thus often only measure effectiveness against moderate to severe disease outcomes.[16] Seroepidemiological studies can detect missed infections by comparing changes in antibody titres, measured in haemagglutination inhibition assay (HAI), pre- and post-season. Traditionally, an HAI titre of 1:40 or greater (seropositivity) in a serological sample has been used to indicate a ∼50% or greater probability of protection against infection for influenza.[17,18] Additionally, a four-fold rise in antibody titre between two time-points in a serological study (seroconversion) has been used to indicate a latent infection in the absence of vaccination.[19,20] However, pre-vaccination HAI titre has consistently been associated with the magnitude of titre boosting post-vaccination or infection.[8,10–13,21] Therefore, the four-fold rise rule could become unreliable under certain circumstances. For example, in an unvaccinated population, where HAI titres are lower, the four-fold rise criterion may be a more reliable method to identify missed infections than in a vaccine-experienced population, which will have higher pre-season titres. In addition, previous seroepidemiological studies have found statistical associations between seroconversion and host factors, including age, infection history, and vaccine history within vaccinated populations.[22–24]

In this study, we develop a flexible Bayesian regression model to estimate the associations and interactions between host factors—pre-vaccination HAI titre, age, sex, vaccine history, and study setting—and individual-level post-vaccination antibody kinetics. We fit the model to a multi-season ongoing cohort study dataset to help understand antibody boosting towards the egg-grown influenza A subtypes vaccine candidate viruses and the cell-grown equivalent viruses, representing circulating viruses. By considering antibody kinetics to vaccinating strains, we use the inferential uncertainty to quantify the importance of each host factor in driving antibody kinetics and assess if the heterogeneity observed among vaccine responses correlates with influenza A subtype kinetics when adjusting for the relevant host factors. For influenza circulating strains, we assess how persistent established heuristics for protection (titre greater than 1:40 and 1:80) and seroconversion (four-fold rise) change post-vaccination to better understand the associations between vaccine responses and protection against infection throughout the season.

## METHODS

### Overview of the cohort study

We consider serological data from an ongoing longitudinal cohort study conducted in Australia.[25] This study commenced recruitment in 2020 and followed an open cohort of healthcare workers (HCW) recruited from hospitals in 6 cities (Brisbane, Newcastle, Sydney, Melbourne, Adelaide, and Perth). HCW could be staff, students or volunteers and were not required to have direct patient contact because vaccination in these hospitals was free to all personnel regardless of risk. Upon enrolment, participants were asked to provide demographic information (sex, age), and influenza vaccination history. Recruited HCW were given a quadrivalent influenza vaccine with a strain composition consistent with the WHO-recommended vaccine strains for that season (See Table S1 for vaccine strains). Sera were collected within two weeks before vaccination and two times after, a mean of 19 (range 8–63) and 166 (range 66–231) days post-vaccination. This analysis used results for 4,958 sera samples collected from 2020-2022, during this period influenza did not circulate in Australia during 2020-2021 [26] and resumed ciruclation in 2022.[27] These 4,958 serum samples came from 1,646 HCW, of which the majority of the HCW (1151/1646) were from frequently vaccinated HCW (vaccinated in 5/5 seasons before enrolment) and 177/1646 were from vaccine-naive HCW (not vaccinated in 5/5 seasons before enrolment). An overview of the study characteristics of this cohort is given in **Table 1**.

**Table 1.**
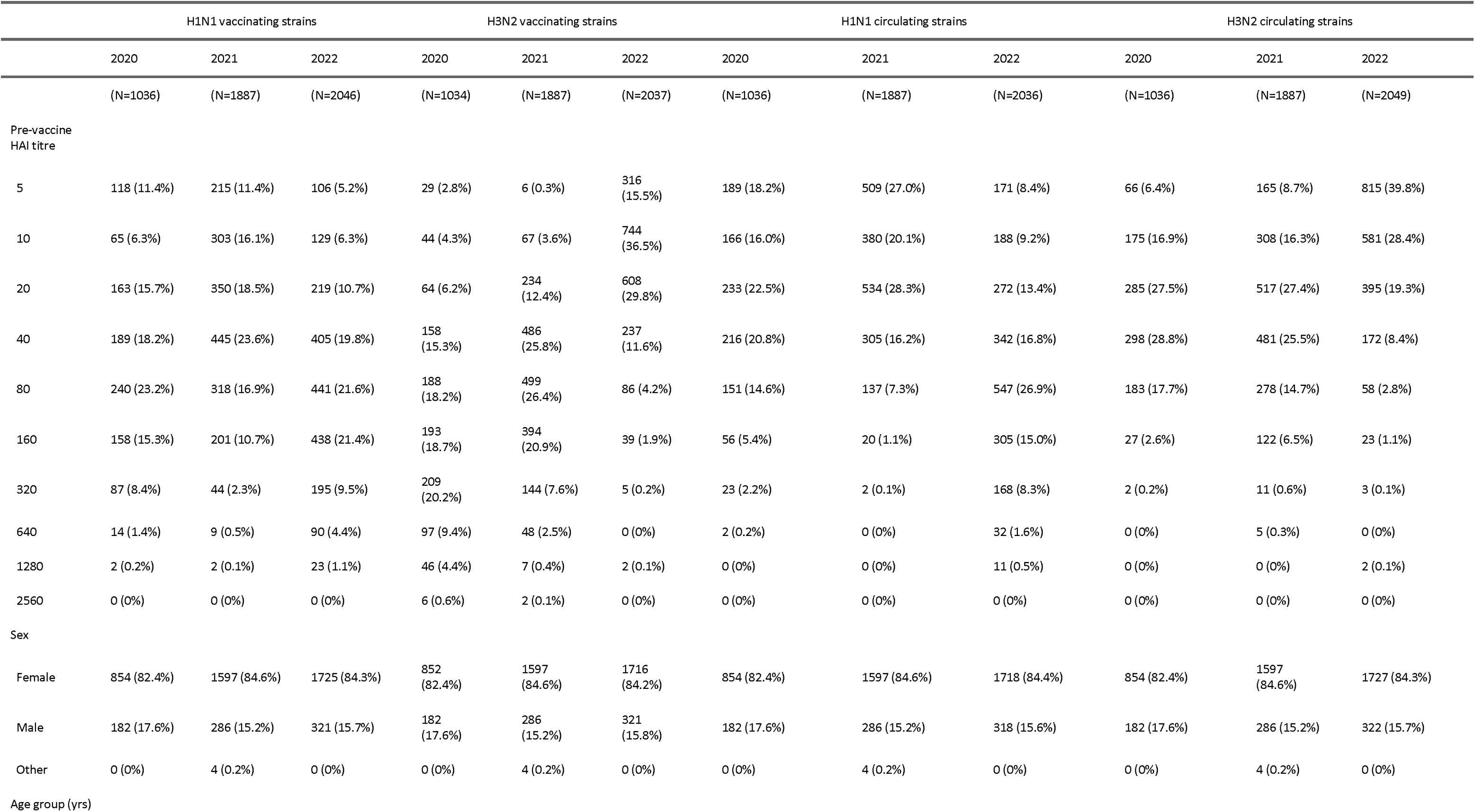
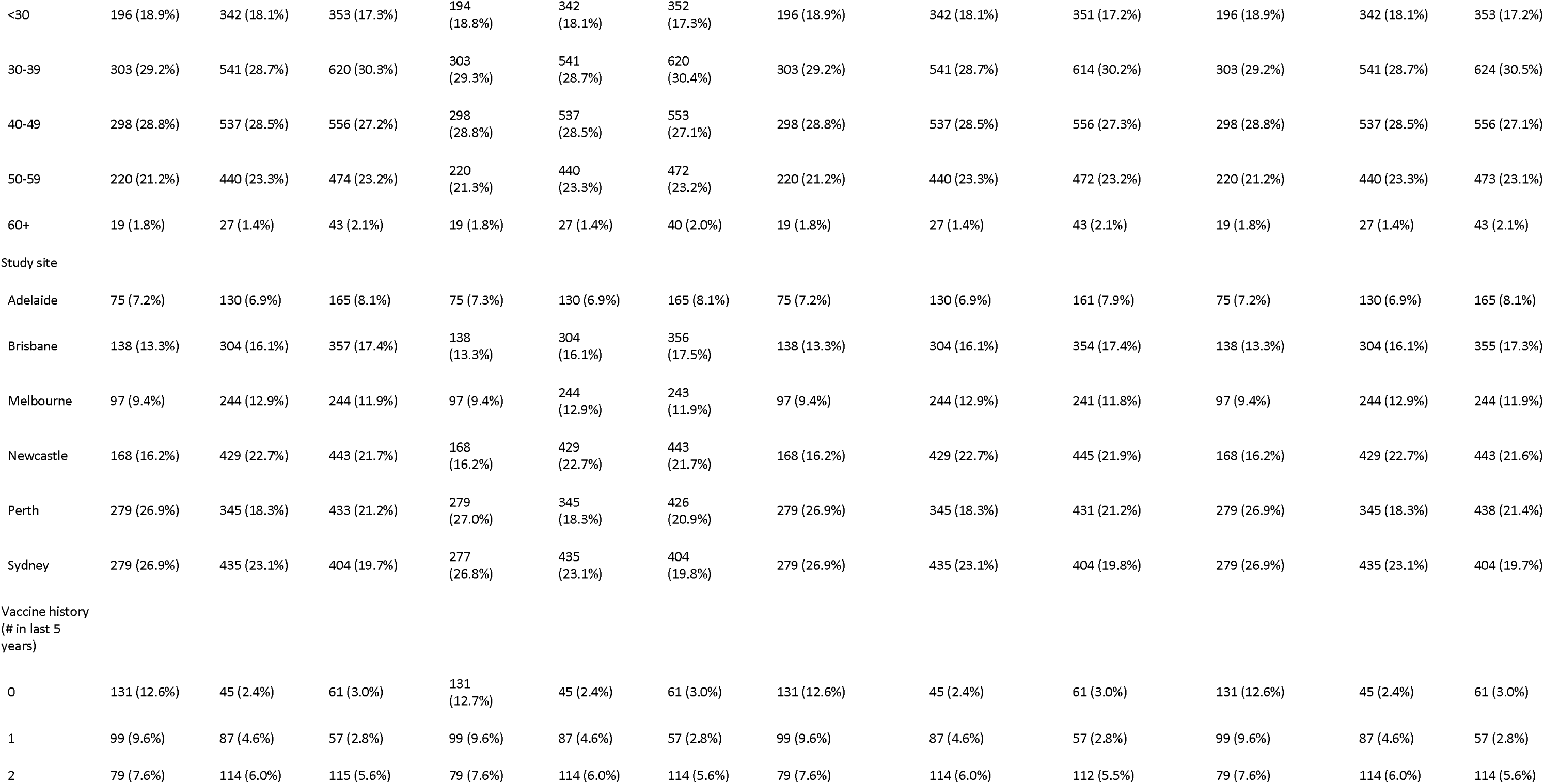
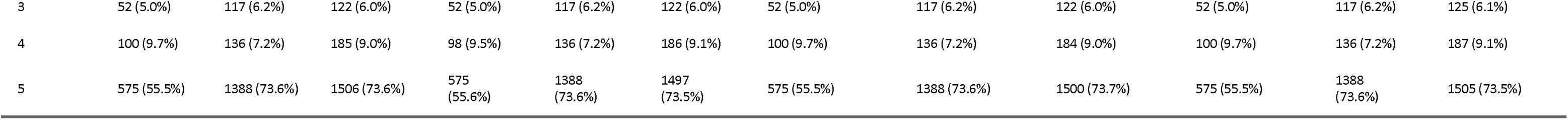
Baseline characteristics of the study populations stratified by covariates for each of the three vaccine types where N is the number of samples.

### Antibody HAI titre

Sera were tested in HAI against relevant egg-grown influenza vaccine strains including wild-type and influenza vaccine reassortant (IVR) strains as follows: A(H1N1) A/Brisbane/02/2018 IVR-190, A/Victoria/2570/2019 IVR-215; A(H3N2) A/South Australia/34/2019, A/Hong Kong/2671/2019 IVR-208, A/Darwin/9/2021. Sera were also tested In HAI against equivalent cell-grown viruses as follows: A(H1N1) A/Brisbane/02/2018, A/Victoria/2570/2019; A(H3N2) A/Darwin/726/2019, A/South Australia/34/2019, A/Darwin/6/2021.

Sera were treated with receptor-destroying enzyme (RDE, Denka Seiken) and adsorbed with a mixture of guinea pig and turkey red blood cells (rbc) to remove non-specific inhibitors and non-specific agglutination. Sera were serially diluted two-fold, ranging from 1:10 to 1:10240. Viruses were diluted to 4 HA units. Assays were performed as previously described (Auladell et al., 2021[28]) using guinea pig red blood cells for A(H3N2) virus titrations and turkey red blood cells for A(H1N1) virus titrations. HI titres were imaged and read using an automated hemagglutination analyser (CypherOne, InDevR) with manual reads for Turkey rbc.

### Description of response and covariates

The covariates for the model were chosen so that they were comparable across all cohorts. These covariates included pre-vaccination HAI titre (10 groups: <1:10, 1:10, 1:20, 1:40, 1:80, 1:160, 1:320, 1:640, 1:1280, >1:2560), age groups (5 groups: <30, 30–39, 40–49, 50–59, 60+ years), gender (3 groups: male, female, other), study site (6 groups: Adelaide, Brisbane, Melbourne, Newcastle, Perth, Sydney ), study year (2020, 2021, and 2022) and the number of influenza vaccines in the last five years (vaccine history) (6 groups: 0, 1, 2, 3, 4, 5). We calculated the individual-level fold-rise in antibody titre relative to the pre-vaccination HAI titre for the response variable at time *t*. The fold-rise in antibody titres was converted to a log, scale throughout the analysis.

### Overview of Bayesian model structure

We developed a Bayesian model that estimates each participant’s antibody response—as measured by HAI titre—at time *t* post-vaccination. The post-vaccine antibody kinetics model assumes an instantaneous boost immediately after vaccination, followed by a linear wane. Two latent parameters describe this model: the peak antibody boosting (log2 scale) and the daily rate of antibody decline (log2 scale). The peak antibody boost was calculated using linear regression with covariates sex, age group, study site, vaccine history, and pre-vaccination HAI titre. Due to the complexity of the immunological mechanisms that relate to pre-vaccination HAI titre and boosting, fitting using a non-parametric form such as a Gaussian Process prior allows for a flexible non-linear association. Further, as the extreme values of the HAI titre values have a low number of samples, a Gaussian process prior will inform these titre values with low power from similar values, giving a more robust estimate than may come from a random-effects model (**see SI methods**). The other covariates were categorical and had a partial pooling effect. We also included the possibility of a product term between pre-vaccination HAI titre and the other covariates to understand the interactions between titre and age/sex/vaccination history. The daily rate of antibody decline also followed a linear regression, but we only considered the impact of pre-vaccination titre values, which we assumed follows a Gaussian process prior. The individual-level effects are added to both the boosting and waning parameters and allow the model to identify unusual individual-level boosting effects that the structure of the population-level model may fail to capture. The model equations, schematics and further details are in **SI Methods.** We fit the Bayesian regression model to four subsets of HAI data from the HCW dataset described below.

To determine which covariates are significantly associated with HAI boosting, we calculated the marginal posterior distribution of peak HAI boosting for covariates: pre-vaccine titre, age group, vaccine history, gender, and site. For a given covariate, if there is a significant difference in the marginal posterior distribution between levels (i.e. 95% credible interval (CrI) does no overlap), then we assumed that the covariate has a significant association with HAI boosting, and we stratified by this covariate in further analyses. We also calculated the marginal posterior distribution for the covariates with interaction terms with pre-vaccination titre (age group, vaccine history, gender, site) to determine the influence of the interaction terms. We considered an interaction term covariate significant if >95% of the marginal posterior distribution is above or below 0 for at least one level of pre-vaccination titre. As we are interested in estimating the mean effects over multiple seasons, we do not stratify by study year, even if the marginal posterior distribution across study year levels is significant. For waning, where we only considered the influence of pre-vaccination HAI titre, we similarly defined the association as significant if the 95% CrI of the marginal posterior distribution between the levels of pre-vaccination HAI titre does not overlap.

### Overview of four influenza viral strains

Using the three years of cohort data, we applied the model to two influenza vaccine viruses and one predominant circulating virus each year to help understand if there are consistent statistical associations between host factors and vaccine-induced HAI boosting for influenza virus strains across different study sites and seasons. First, to understand the vaccine-induced antibody kinetics response to vaccination strains, we fit the model to HAI titre to A(H3N2) egg-grown vaccine viruses A/South Australia/34/2019, A/Hong Kong/2671/2019, and A/Darwin/09/2021 for 2020, 2021 and 2022 respectively (A(H3N2) vaccinating strains). Similarly, for A(H1N1), we fit the model with HAI titres to egg-grown strains; A/Brisbane/02/2018, A/Victoria/2570/2019, A/Victoria/2570/2019 for 2020, 2021 and 2022 respectively (A(H1N1) vaccinating strains). To help understand how egg-grown adaptations impact vaccine-induced antibody kinetics, we also fit the model to HAI titres for cell-grown viruses. For this, cell-grown A(H3N2) viruses, matching the same clade as the seasons’ egg-grown vaccine strains for 2020, 2021 and 2022 were used, namely A/South Australia/34/2019 (3C.2a.1b.2), A/Darwin/726/2019 (3C.2a.1b.1b), A/Darwin/6/2021 (3c2a1b.2a.2) respectively (A(H3N2) circulating strains) and for cell-grown A(H1N1) viruses, we used A/Brisbane/02/2018, A/Victoria/2570/2019, and A/Victoria/2570/2019 for 2020, 2021 and 2022 respectively.

### Implementation

All data cleaning, posterior inference, and plotting were performed in R (v.4.0.1) through VScode. The fitting of the Bayesian regression model was sampled through Hamiltonian Monte Carlo via cmdstanr (v. 0.5.3). A GitHub repository with all the code needed to reproduce this work is given at https://github.com/dchodge/ab_boosting_published. Data for the study is available upon request from the corresponding author.

### Model Validation

To assess the fit of the Bayesian model, we compare the model-fitted HAI titre boost on the day of bleed with the HAI titre data used to fit the model. We found for all four groups of viruses (A(H1N1) vaccine, A(H3N2) vaccine, A(H1N1) circulating, and A(H3N2) circulating), that the HAI titre boosts values were well correlated by the model fitted values with 99.4%, 99.2%, 99.1% and 99.2% of model estimates within a one-fold-change unit of the data for strain types respectively (**SI Figure 1)**. If the fold-rise in the data exceeded 16, the model consistently underestimated the HAI titre boosting. However, titre fold-rises of this magnitude were uncommon, with 6.5%, 2.6%, 1.6% and 1.9% of samples seeing a 32-fold rise in titre or higher for the three influenza strain types.

### Ethics Approval

The Royal Melbourne Hospital Human Research Ethics Committee of Royal Melbourne Hospital gave ethical approval for study protocol and protocol addendums for follow-up of COVID-19 vaccinations and SARS-CoV-2 infections (HREC/54245/MH-2019). LSHTM Observational Research Ethics Committee of London School of Hygiene and Tropical Medicine gave ethical approval for the use of this data for analysis (ref 22631).

## RESULTS

### Determining covariates that significantly influence antibody-boosting

For A(H1N1) vaccinating strains, our estimates suggest that from the covariates considered, only pre-vaccination HAI titre and vaccine history significantly influence subsequent HAI boosting (Figure 1, SI *Figure 2 for posterior distributions of regression coefficients SI Figure 3 for marginal posterior distributions for all covariates*) (**Figure 1A**). At pre-vaccination HAI titres of <1:10, 1:10, and 1:20, titres were predicted to rise 19.0-fold, 12.2-fold and 6.9-fold with Posterior Predictive Intervals (PPI) of 15.0–26.9, 10.1–16.0, and 5.8–8.8, respectively (**Figure 1A**). We also found that the expected titres did not rise above the four-fold threshold when pre-vaccination titres were 1:80 or higher. For vaccination history, titres were predicted to rise 8.2 (95% PPI 6.8–10.5) fold in the absence of prior vaccination compared to a 4.2 (95% PPI 3.6–5.4) fold if vaccinated with two or more vaccines in the last five years (**Figure 1B).** There was weak evidence of an effect of interaction between pre-vaccination titre and vaccination history, suggesting that boosting remains high for infrequently vaccinated individuals independently of pre-vaccination HAI titre (**Figure 1B**). We also find that the waning rate of HAI titre is significantly associated with pre-vaccination HAI titre, with lower HAI titres experiencing faster waning rates (**Figure 1A**).

**Figure 1.**
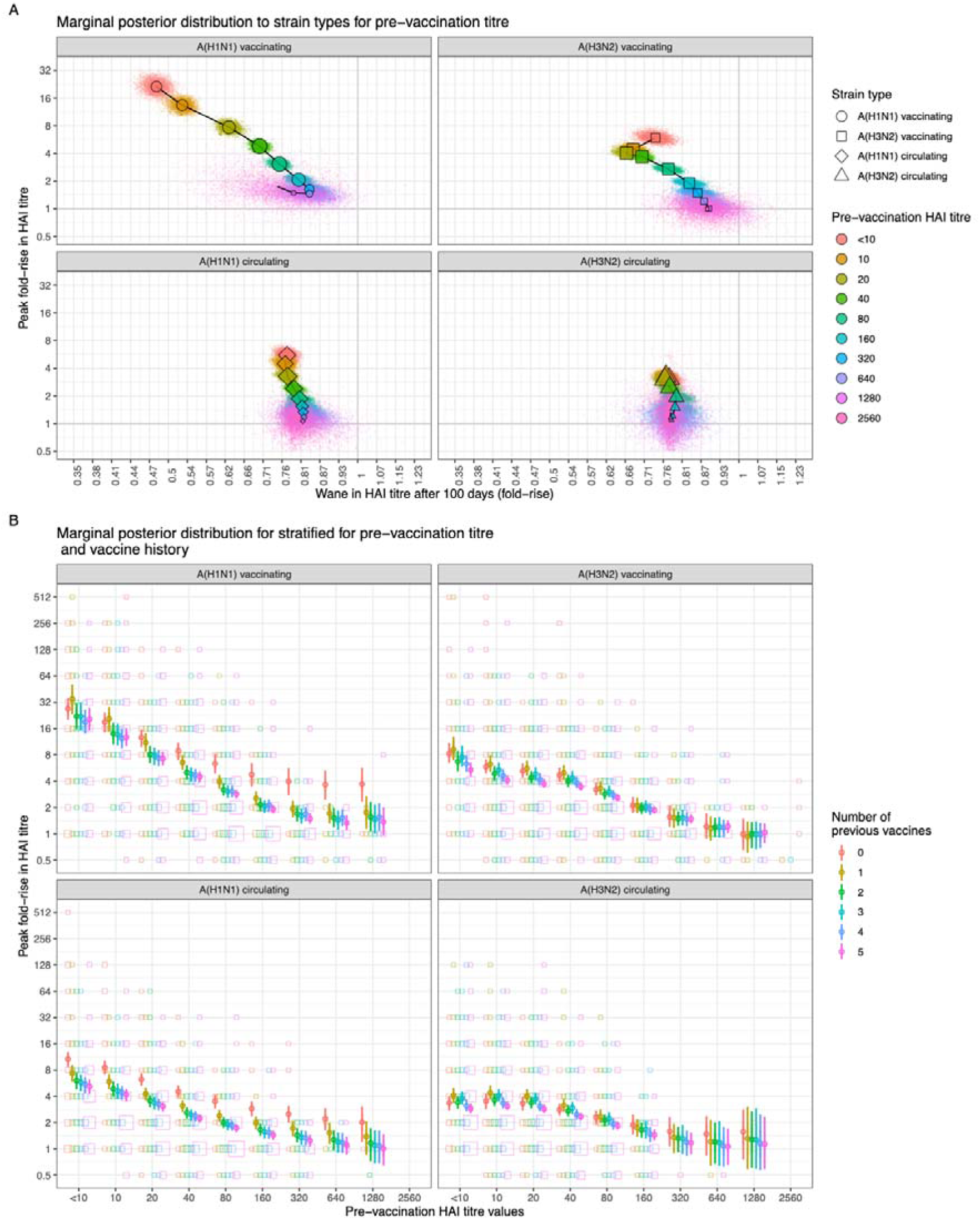
Bayesian marginal posterior distribution for covariates as a measure of inferential uncertainty. (A) The marginal posterior distributions of boosting and waning stratified by pre-vaccination HAI titre for the four virus strains. The point markers are the mean value of the posterior, and the smaller point marker represents a posterior sample. (B) The marginal posterior distribution of boosting stratified by pre-vaccination HAI titre and the number of previous vaccines for the four virus strains. The point marker indicates the posterior mean and the uncertainty represents a 50% and 95% posterior predictive interval (thick and line respectively).

Pre-vaccination HAI titre and vaccine history also strongly influence HAI boosting for A(H3N2) vaccinating strains (see *SI Figure 4 for posterior distributions of regression coefficients and SI Figure 5 for marginal posterior distributions for all covariates*). Lower HAI pre-vaccination titres are associated with higher boosting, with a pre-vaccination HAI titre of <1:10, 1:10, and 1:20 predicted to rise an estimated 6.1 (95% PPI 5.2–7.5), 4.4 (95% PPI 4.0–5.2), and 4.1 (95% PPI 3.7–4.7) fold respectively (**Figure 1A**). Titres did not rise above the four-fold threshold when pre-vaccination HAI titres were 1:40 or higher. For vaccination history, we find strong evidence of an effect of interaction between pre-vaccination titre and vaccination history, suggesting that the magnitude of the difference in boosting with increasing years of prior vaccination diminishes as pre-vaccination titre increases (**Figure 1B)**. We estimate that for HCW who had pre-vaccination titres of <1:10 those with no vaccinations in the last five years have an 8.5 (95% PPI 7.0–11.1) fold boost, compared to a 5.6 (95% PPI 4.7–6.8) fold boost for those who had five vaccinations in the last five years (**Figure 1B**). We also find that the waning rate of HAI titre is significantly associated with pre-vaccination HAI titre, with lower HAI titres seeing faster waning rates (Figure 1A).

For A(H1N1) and A(H3N2) circulating strains, we also find pre-vaccination HAI titre and vaccine history strongly influence HAI boosting (**Figure 1A**, see *SI Figure 6 and 8 for posterior distributions of regression coefficients and SI Figure 7 and 9 for marginal posterior distributions for all covariates*). We find boosting was attenuated for circulating compared to vaccines strains and failed to reach a four-fold for nearly all pre-vaccination titres. We also find the waning of HAI titre is not significantly influenced by the pre-vaccination HAI titre (**Figure 1A**). For all strain types in **Figure 1B**, we plot the outcome variability of the underlying data (squares) to highlight the large variability in outcome compared to the inferential uncertainty.

We find large differences in peak HAI boosting between seasons across all three vaccine types. For A(H1N1) vaccinating strains, HAI boosting was significantly higher in 2021 than in other years (8.1-fold vs. 3.2 and 3.1). For A(H3N2) vaccinating strains, we find 2022 has a significantly higher boosting than 2020–2021 (4.1 vs. 2.6 and 2.7) with a similar observed trend for A(H3N2) circulating strains, albeit with attenuated boosting compared to A(H3N2) vaccinating strains (**SI Figures 2, 4, 6, 8**).

### Relating aggregated antibody kinetics to seroconversion and protection

Using our estimates of antibody boosting and waning, we generated predictions for antibody trajectories aggregated across different levels pre-vaccination titre and vaccine history calculated for up to 220 days post-vaccination for each strain type (**Figure 2)**. To understand how these kinetics influence population-level immunity, we estimate the posterior predictive interval of the duration of i) seroconversion (a 4-fold rise from their pre-vaccination HAI titre) and ii) seroprotection (HAI titre equal to or exceeding 1:40, which is associated with a 50% reduction in risk of infection or 1:80 associated with a 90% reduction in risk of infection[18,29]) for influenza A subtype circulating strains For A(H1N1) circulating strains, we find the duration of seroconversion for those with pre-vaccination titres of <1:10, 1:10, and 1:20 is 308 (95% PPI 229–389), 217 (95% PPI 151–284), and 100 (95% PPI 42–155) days for individuals with less than two vaccines in the last five years (infrequently vaccinated) and 130 (95% PPI 52–198), 46 (95% PPI 0–101), and 0 (95% PPI 0–0) days for individuals with two or more vaccines in the last five years respectively (**Figure 3A**). For A(H3N2) circulating strains, the duration of seroconversion has a mean of between 0–10 days for all vaccine histories and pre-vaccination titres. For seroprotection, assuming a threshold value of 1:40, infrequently vaccinated individuals are protected 171 (95% PPI 128–220) and 159 (95% PPI 120–200) days longer compared to frequently vaccinating individuals for A(H1N1) circulating viruses with pre-vaccination titers of 1:10 and 1:20 respectively (**Figure 3B**). At a seroprotection threshold of 1:80, we similarly see the benefit of being infrequently vaccinated, providing 192 (95% PPI 145–244) days longer seroprotection assuming a pre-vaccination titre of 1:40. For A(H3N2) circulating viruses, there is a slightly longer duration of protection for infrequently vaccinated individuals of around 50 days, however these beneficial effects are limited to the those with an HAI titre value of 1:20 (at a 1:40 threshold) or 1:40 (n at a 1:80 threshold).

**Figure 2.**
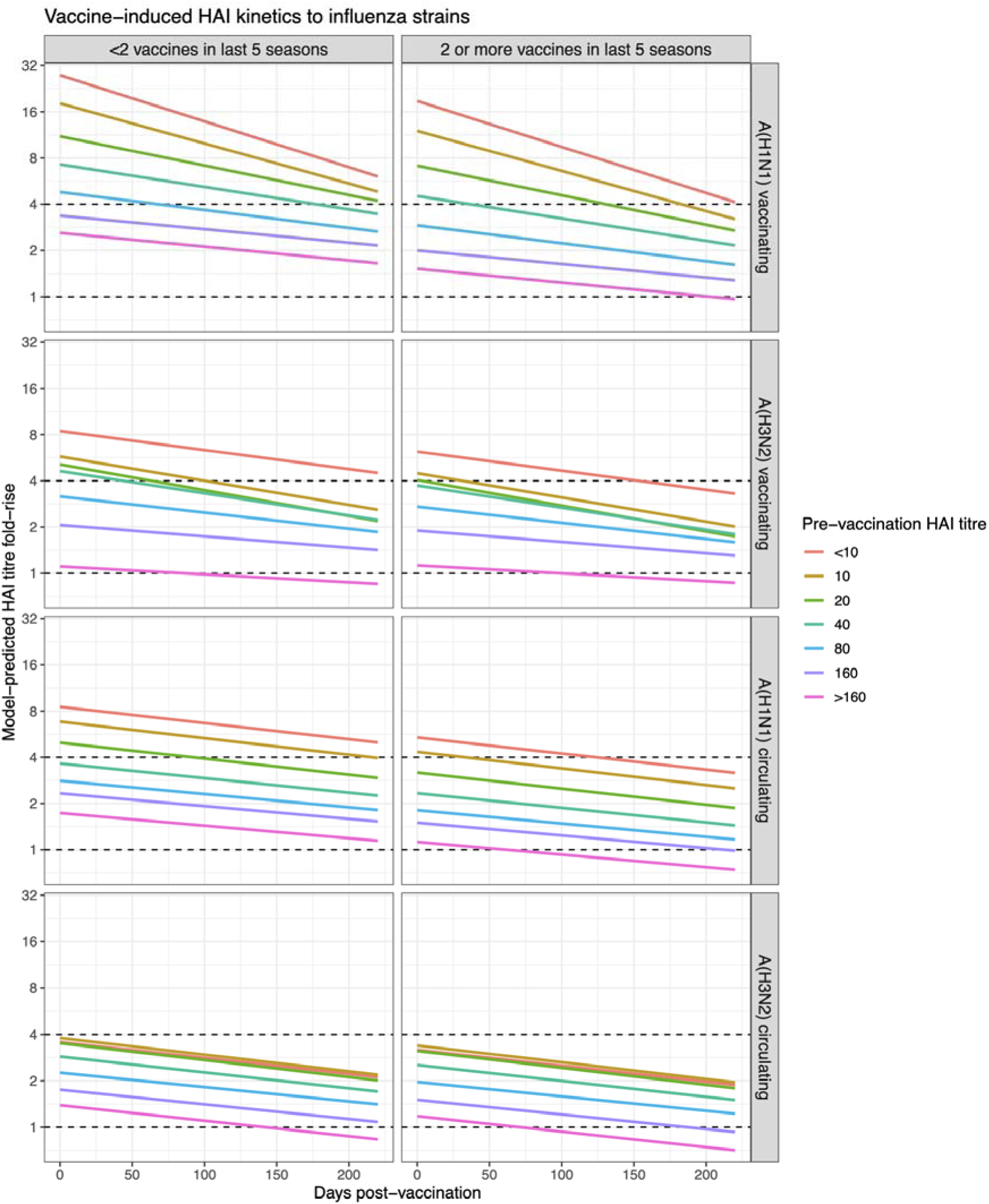
Expectation of post-vaccination kinetics from marginal posterior distributions of latent parameters. The mean post-vaccination HAI boosting (fold-rise) trajectories to virus strains stratified by pre-vaccination HAI titre and vaccination history (infrequently vaccinated and frequently vaccinated).

**Figure 3.**
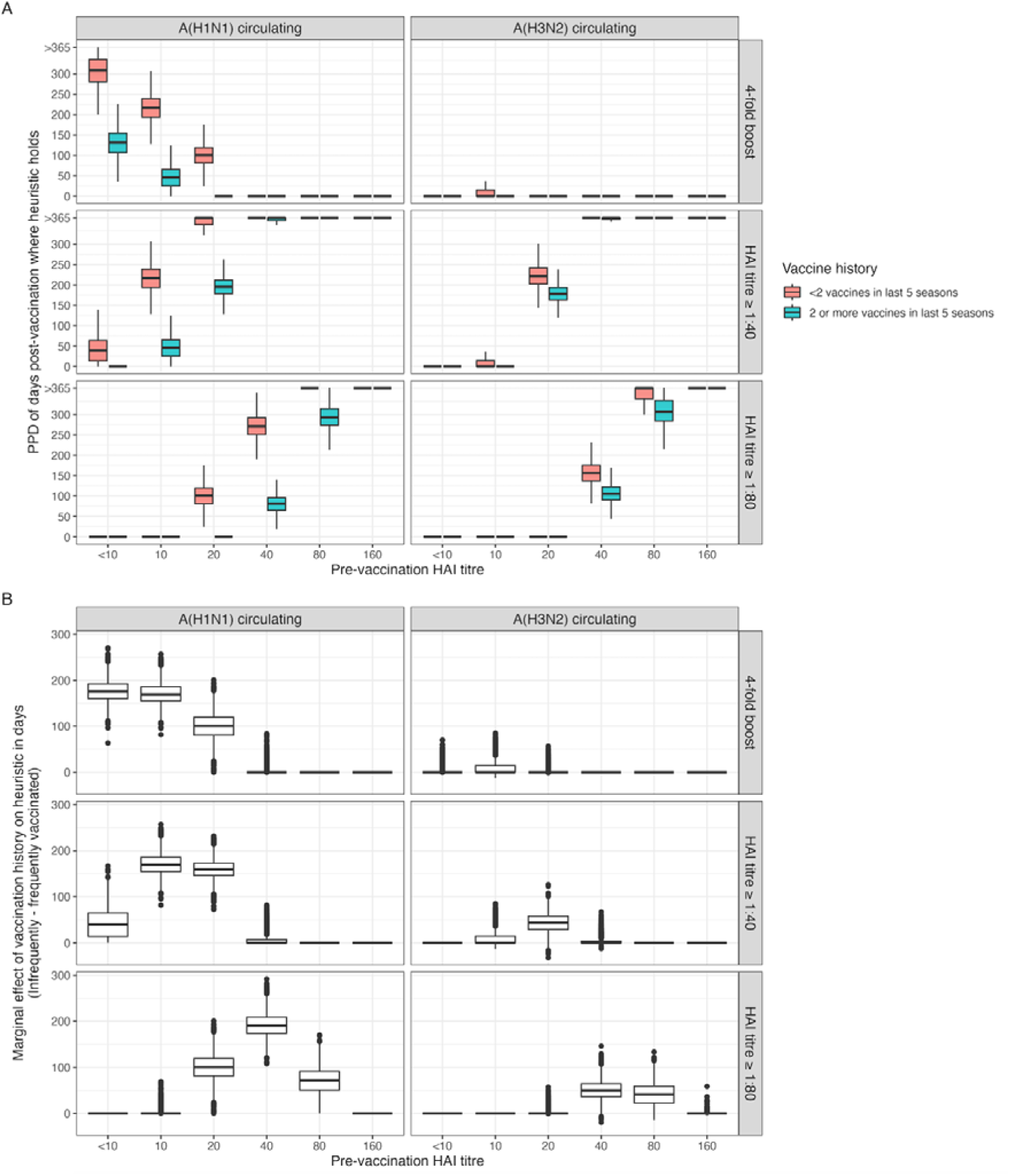
Post-vaccination duration of seroconversion and sreoprotection from inferred statistical model. (A) Posterior predictive distribution (PPD) for the duration that titre rise is ≥4-fold and that HAI titre is ≥1:40 and ≥1:80 after vaccination stratified by vaccination history and pre-vaccination titre. (B) The PPD for the differences in days that titre rise ≥4-fold and that HAI titre is ≥1:40 and ≥1:80 between the two vaccine histories.

### The practical significance of the inferred statistical model

The latent antibody trajectories aggregated across covariate levels of pre-vaccination titre and infection history from the models suggest significant differences in the duration of seroprotection and seroconversion. However, the large observed individual-level outcome variability means it’s difficult to ascertain if these aggregated differences in antibody kinetics translate to a meaningful difference in protection when we consider individual-level variation across a whole cohort. To assess this, we use the individual-level fits from the regression model and determine how long each individual’s post-vaccination HAI titre remains high enough to be four-fold higher than the pre-vaccination HAI titre, i.e. seroconverts and/or is seroprotected from infection **(Figure 4**). We find that the mean value of the duration of seroconversion and seroprotection is higher in the infrequently vaccinated cohort across nearly all pre-vaccination HAI titre values (**Figure 4A**). For A(H1N1) circulating strains, we find a significant difference (according to a Kolmogorov-Smirnov test) between the empirical distributions for the individual-level duration of protection and seroconversion between frequently and infrequently vaccinated populations across most pre-vaccination titre levels. The magnitude of this significant difference according to a Cohen’s D effect size suggests that the magnitude of the difference is large (above 0.8) for pre-vaccination HAI titres less than or equal to 1:40 (**Figure 4B**). For A(H3N2) circulating strains, there is no significant difference between the individual-level empirical distributions between frequently and infrequently vaccinated individuals for most pre-vaccination HAI titres, suggesting that vaccine history has little practical significance on seroconversion rates. For seroprotection with A(H3N2) circulating strains, significant differences in the empirical distributions and in the magnitude of the effect size are seen between infrequently and frequently vaccinated for those with pre-vaccination titres of 1:40 and 1:80 only.

**Figure 4.**
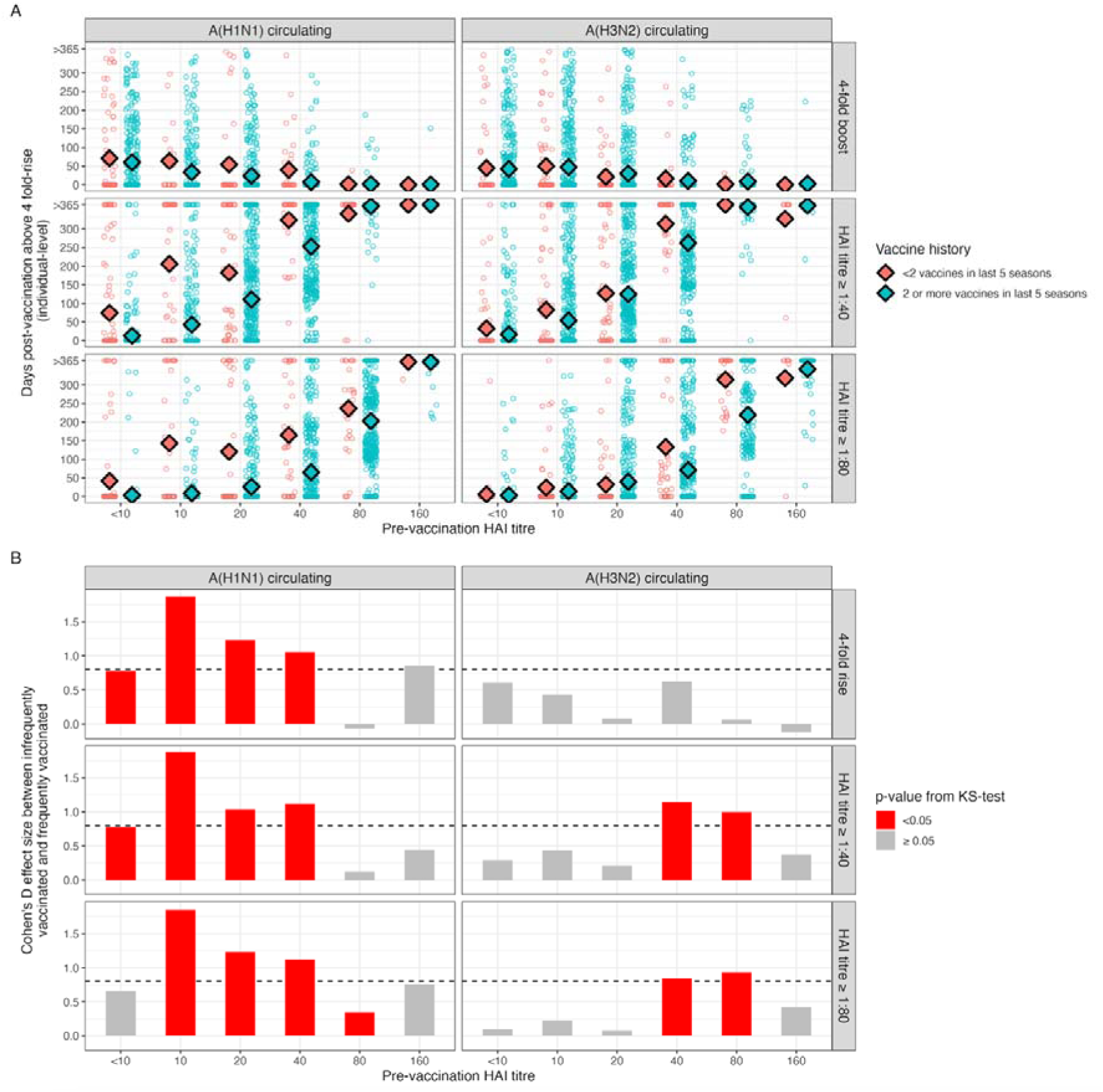
Post-vaccination duration of seroconversion and protection when considering outcome variability. (A) Individual-level estimates for the duration that titre rise is ≥4-fold and that HAI titre is ≥1:40 and ≥1:80 after vaccination stratified by vaccination history and pre-vaccination titre. Large diamond point marker individuals mean of sample. (B) Effect size from Cohen’s D and the p-value from a Kolmogorov-Smirnoff test to assess the significance of the difference between the two vaccine history levels for the duration post-vaccination above a threshold heuristic.

Comparing the posterior predictive distributions of individual-level dynamics between strain types between individuals, we see that boosting and waning estimates between A(H3N2) and A(H1N1) vaccinating strains are not strongly correlated, but boosting to H1N1 is significantly higher. Boosting and waning between the A(H3N2) vaccinating and A(H3N2) circulating strains are slightly correlated, though A(H3N2) circulating strain boosts are generally attenuated (**SI Figure 8).**

## DISCUSSION

We found that pre-vaccination HAI titre and vaccination history significantly influenced HAI antibody boosting induced by vaccination. This result holds for antibodies against both A(H1N1) and A(H3N2) vaccination strains, as well as against cell-grown circulating strains. We used model-predicted antibody trajectories to estimate the expected duration of seroprotection (HAI titre ≥1:40) and seroconversion (≥4-fold rise) for cell-grown circulating strains and found that those with lower pre-vaccination HAI titres experience longer durations than those with higher pre-vaccination titres. In addition, for A(H1N1) circulating strains, our statistical models suggest that infrequently vaccinated populations can experience a mean of 50-300 days longer duration of seroprotection against influenza compared to frequently vaccinated populations if their pre-vaccination titre is below 1:40. These observations result in practically significant differences in the duration of seroprotection when we consider individual-level variation across a whole cohort for circulating strains. However, for circulating A(H3N2) strains, the attenuation in boosting compared to vaccinating strains means that there is almost no seroconversion observed, and vaccine-induced seroprotection is only afforded to those individuals with pre-vaccination HAI titres close to the protection thresholds.

Our observation that pre-vaccination titres significantly influence HAI antibody boosting is consistent with previous seroepidemiological studies.[8,10–13,21,30] These studies suggest individuals with lower pre-vaccination titres see higher fold rises (e.g. 16 fold for <1:10[21]) compared to those with higher pre-vaccination titres, with four-fold rises becoming uncommon after pre-vaccination titres of 1:40/1:80. [10,21,30] This trend is consistent across influenza subtypes and between HAI titres and micro-neutralisation assays. Previous models which fit individual-level antibody trajectories as a latent parameter have also considered the influence of pre-exposure HAI titres on HAI boosting. Ranjeva et al.[31] used individual-level models of post-infection antibody trajectories and found they were improved by assuming that (i) boosting decreases with increasing pre-infection titre and (ii) that there is limited seroconversion when pre-infection titres exceed 1:80. Similarly, a model by Hay et al.[32], which builds antibody trajectories given different sequences of influenza exposure types in ferrets, found that pre-vaccination HAI titre-dependent boosting leads to better model fits than without. The framework in this study augments the titre-dependent boosting mechanisms outlined in these previous models in two ways. First, instead of a linear relationship, we consider a flexible non-parametric relationship between log pre-vaccination titre and log fold-rise, allowing for more complex dynamics relating the two to be described. Our fitted relationship suggests an almost linear decrease in log fold-rise as log pre-vaccination HAI titre increases until a pre-vaccination titre of 1:640, after which there is no fold-rise. The second way we extend previous models, considering the influence of pre-vaccination titre, is by incorporating a hierarchical Bayesian regression structure, allowing for the quantification of age, sex, and vaccination history on vaccination-induced antibody kinetics. Therefore, after calculating the marginal distributions, we reduce potential confounding and allow for more accurate descriptions of covariate-specific antibody trajectories.

The influence of vaccination history on post-vaccination antibody kinetics has also been observed in previous seroepidemiological studies. [23,33–35] These studies found that individuals recently vaccinated with influenza experience attenuated boosting compared to those without recent vaccination. However, these studies often report higher pre-vaccination HAI titres, suggesting that the attenuated boost attributable to vaccination history might be confounded by attenuation in boosting due to pre-vaccination titre.[35] We, therefore, focus comparisons to our study with studies which explicitly consider the influence of pre-vaccination HAI titre and the impact of vaccine history on antibody kinetics.[8,36] Beyer et al.[36] used 1,119 paired serum samples from 681 individuals to estimate post-vaccination HAI titre in a multiple linear regression model. Across all dependent variables (pre-vaccination titre, vaccine, age, and gender), they found that increasing pre-vaccination titre and vaccination history had a significant negative effect on post-vaccination HAI titre boosting. In contrast, age and gender did not significantly influence titre boosting across influenza subtypes. Wu et al.[8] used samples from 1,300 individuals and fit a multiple linear regression model to predict seroconversion. They found that increasing pre-vaccination HAI titre and vaccination history were the two most important predictive covariates for seroconversion rates, with age, body mass index (BMI), sex, race, and comorbidities having little importance.[21,30,31] These observations align with the results of our models; however, we extended these frequentist linear models by implementing a Bayesian framework with partial pool effects across covariates and allowing for interaction terms between pre-vaccination titre and other covariates. We found that vaccination history significantly influences boosting, but only as an interaction term with pre-vaccination titre for A(H3N2) vaccinating strains. Therefore, the degree of influence vaccination history has on titre-boosting changes depending on pre-vaccination HAI titre. This observation could explain the variable impact that vaccination history has on titre boosting, particularly in studies which do not account for pre-vaccination HAI titre.[23,33,34]

A limitation of this model is that the post-vaccination antibody kinetic trajectories are a simple representation of key dynamics (a variable boost with variable linear wane) compared to previous modelling efforts, which use piecewise linear or exponential functions.[31,32,37] We chose this kinetics structure for several reasons. First, it reduces the number of parameters needed in the hierarchical model structure simplifying the fitting process. The latent parameters, peak titre boosting and wane rate per day are also easily interpretable and don’t require complex post-processing to find informative metrics. Second, it would be challenging to infer these complex individual-level trajectories robustly, given the dataset, as bleed dates are constrained to values around 14- and 180-days post-vaccination. Cohort studies with multiple bleeds for the first few weeks following vaccination would help better inform complex initial antibody kinetics. The longer-term dynamics of antibody kinetics suggest a plateauing effect to a set point.[18,37] As we infer a linear structure in our post-vaccination kinetics, we only infer trajectories up to 365 days post-vaccination to help prevent incorrectly estimating the duration of protection and seroconversion at long time scales. Future models could extend this framework by using more complex antibody kinetics structures, such as in-host B-cell kinetics and multiple antibody production sites (e.g. plasmablasts and plasma cells). This can be done implicitly by assuming antibody kinetics follow power function decay functions[38] or by explicitly modelling antibody secretion using systems of Ordinary Differential Equations (ODEs) which relate antigen-secreting cells and antibody titres.[39]

The statistical model without an individual-level variation model fails to explain the outcome variability in the underlying data, meaning predicting individual patterns of HAI boosting using pre-vaccination titre and vaccine history remains challenging. There are augmentations to the model which could help explain some existing outcome variability: most notable is the effect of prior influenza of infection on antibody kinetics.[28] This was not included in this model because there was a lack of information on infection history before 2020. (It is worth noting that there was little influenza virus circulation in Australia between 2020–2021 due to the COVID-19 restrictions, so infection rates in our cohort during that period are likely nil.) The lack of infection history combined with unusual influenza dynamics over this period may help explain some of the outcome variability the model failed to capture. However, there are processes which drive outcome variability that this model cannot consider. This includes variability from other in-host immune processes, such as innate immunity, cellular immunity, genetic polymorphism and epigenetic factors.[40,41]

This study finds that the circulating influenza A strains that differ antigenically from the egg-grown vaccinating strains have notably muted HAI boosting when stratified by pre-vaccination titre and vaccine history. The degree of this attenuation in boosting is so great for A(H3N2) that the influence of vaccine history no longer results in significant differences in outcome variability for many pre-vaccination HAI titres. The observed differences in practical significance in seroconversion between the vaccination strains and circulating strains could explain the seasonal variability of the influence of vaccination history on observed vaccine effectiveness.[42,43] Future model development will incorporate the influence of the antigenic distance between successive vaccinating and circulating strains into the antibody kinetics framework. This could be done by using existing metrics of antigenic advance or considering the influence of vaccination on boosting HAI titres across various strain landscapes.[44] This would allow the exploration of the antigenic distance hypothesis, which suggests that the efficacy of vaccines is muted in a vaccinated individual if a circulating strain is antigenically distant from the previous two antigenically similar vaccine strains.[45]

This study provides robust statistical inference on the host-specific driving forces behind influenza HAI titre kinetics within a single Bayesian framework. This flexible Bayesian model provides the quantifiable non-parametric relationship between pre-vaccination HAI titre and prior vaccination history and established heuristics such as seroconversion and protection. After accounting for pre-vaccination titre, we show that vaccination history significantly influences the duration of seroconversion and seroprotection against influenza, both with the statistical framework and within the outcome variability with individual variation. Future studies that rely on serological and antibody kinetics endpoints should include these two essential covariates to ensure accurate epidemiological inference.

## Data Availability

https://github.com/dchodge/ab_boosting_published. However, some data needs to be requested from the corresponding author.

## COI

David: None

Annette Fox: reports receiving study funding from Sanofi and payments to her research group from Evidera Inc for consulting work.

Stephany: None

Louise: None

Yi: None

Jessica: None

Sheena Sullivan: Reports consulting for CSL Seqirus, Moderna, Pfizer, and Evo Health.

Adam Kucharski: None

## Author contributions

DH: Conceived and designed the study. Performed the formal analysis and development of the methodology and software. Prepared and created the visualisation and wrote, reviewed and edited the manuscript.

SSO: Involved in the preparation and analysis of biological as well as data collection, analysis, and curation.

LC: Involved in the preparation and analysis of biological as well as data collection, analysis, and curation

YL: Involved in the preparation and analysis of biological as well as data collection, analysis, and curation.

JH: Involved in the preparation and analysis of biological as well as data collection, analysis, and curation.

AF: Conceived the study and was involved in the preparation and analysis of biological as well as data collection, analysis, and curation. Reviewed and edited the manuscript.

SS: Conceptualisation and supervision. Reviewed and edited the manuscript.

AK: Conceptualisation and supervision. Reviewed and edited the manuscript.

## FUNDING

This work was supported by the US National Institutes of Health (grant # R01AI41534, SGS, SF, AJK) and by the US Centers for Disease Control and Prevention (contract #HHSD2002013M53890B and #NMR-9619/CDC13FED1310208/NMR9864/CDC16FED1612328, AJK). AJK was also supported by a Sir Henry Dale Fellowship jointly funded by the Wellcome Trust and the Royal Society (grant Number 206250/Z/17/Z).

## Notes

### Summary of Updates

Added in analysis on H1N1 circulating strains and rephrased discussion.

