## Appendix for "Quantifying the impact of pre-vaccination titre and vaccination history on influenza vaccine immunogenicity"

Supplementary Methods to “Post-vaccine HAI antibody kinetics are driven by pre-vaccination HAI titre and vaccine history”

”

1. METHODS: Description and justification of Bayesian model structure

This supplementary provides a detailed description of the Bayesian regression hierarchical model structure for the antibody kinetics model described in the manuscript. Biological and virological considerations, in-host immune dynamics and experimental measurement error of HAI assays have informed the structure of the regression model. Further, we provide casual arguments for including the relevant covariates in the regression equation and a biological justification of the likelihood function used. A schematic showing an overview of the Bayesian model is given in **Figure SM1.**

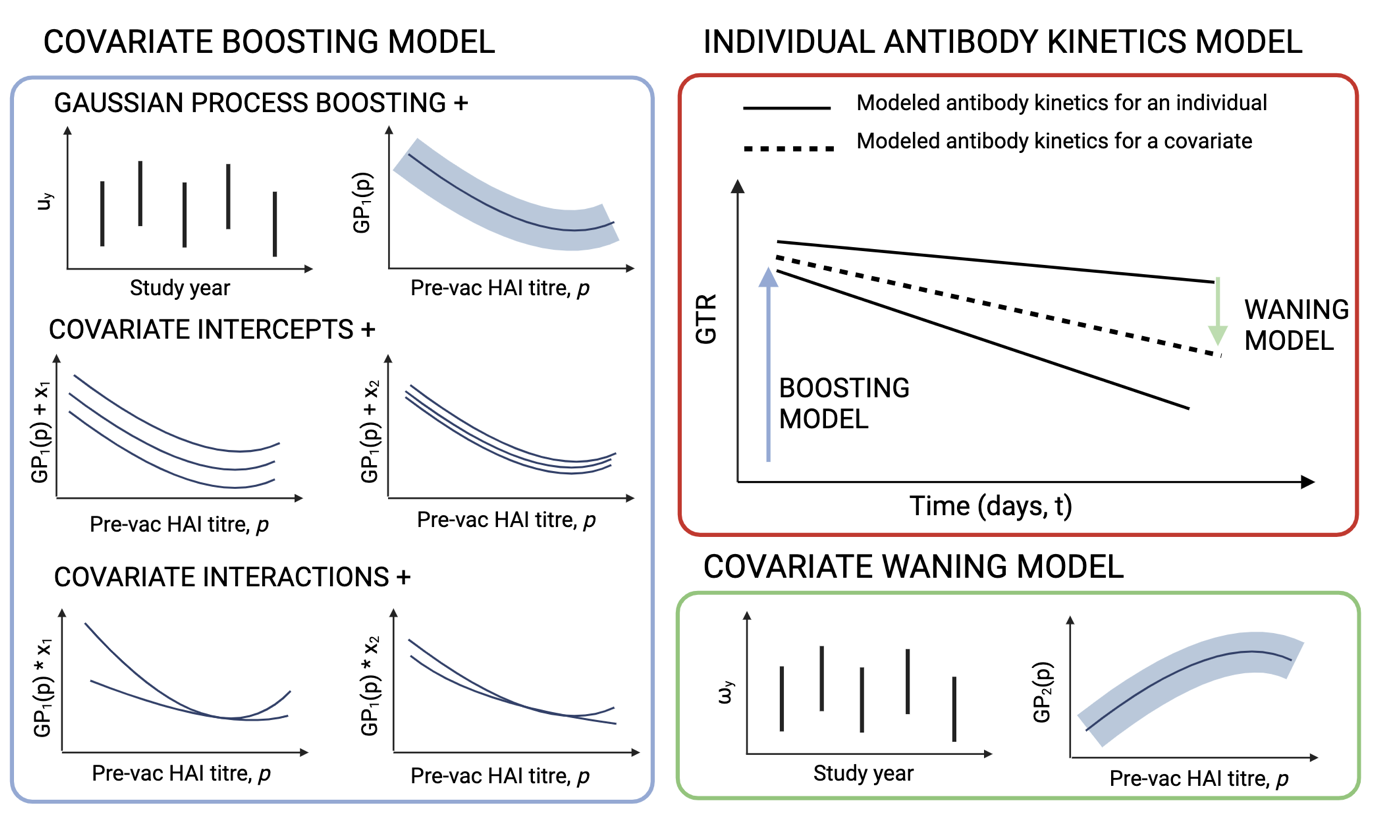

**Figure SM1.** Schematic showing how the various components of the model structure relate (covariate boosting, covariate waning, and individual-level antibody kinetics). *GP* refers to a Gaussian process prior.

1.1 Overview of the dataset

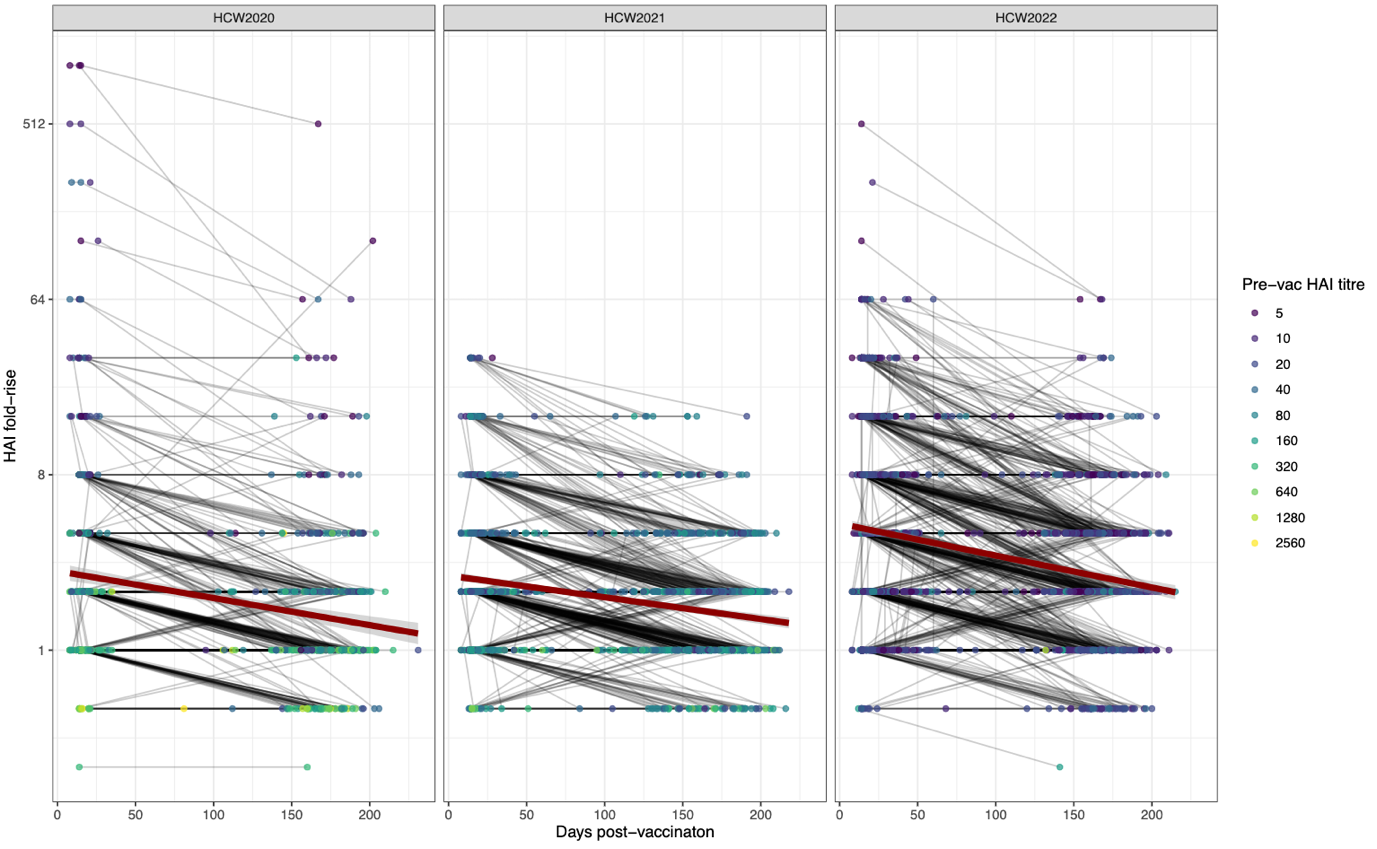

**Figure SM2.** Overview of post-vaccination HAI titre boosting for each study year.

1.2 Description of model variables
    1.2.1 The HAI assay

HAI assays measure the highest 2-fold dilution of serum that prevents influenza-induced hemagglutination of erythrocytes. Consequently, the HAI titres values are discrete and on an exponential scale. In this study, the fold dilutions of the HAI titre (1:h_raw_) are <1:10, 1:10, 1:20, 1:40, 1:80, 1:160, 1:320, 1:640, 1:1280, and 1:2560, with higher titres referring to a higher concentration of influenza antibodies (see (**Figure SM3**). We linearise these serum dilution values through the formula x_i_ = log_2_(h_raw_ / 5) and <1:10 = 0 (**Table SM1).** We call the *log_2_ HAI titre* the HAI titre value of an individual on the scaled log_2_ dilution scale.

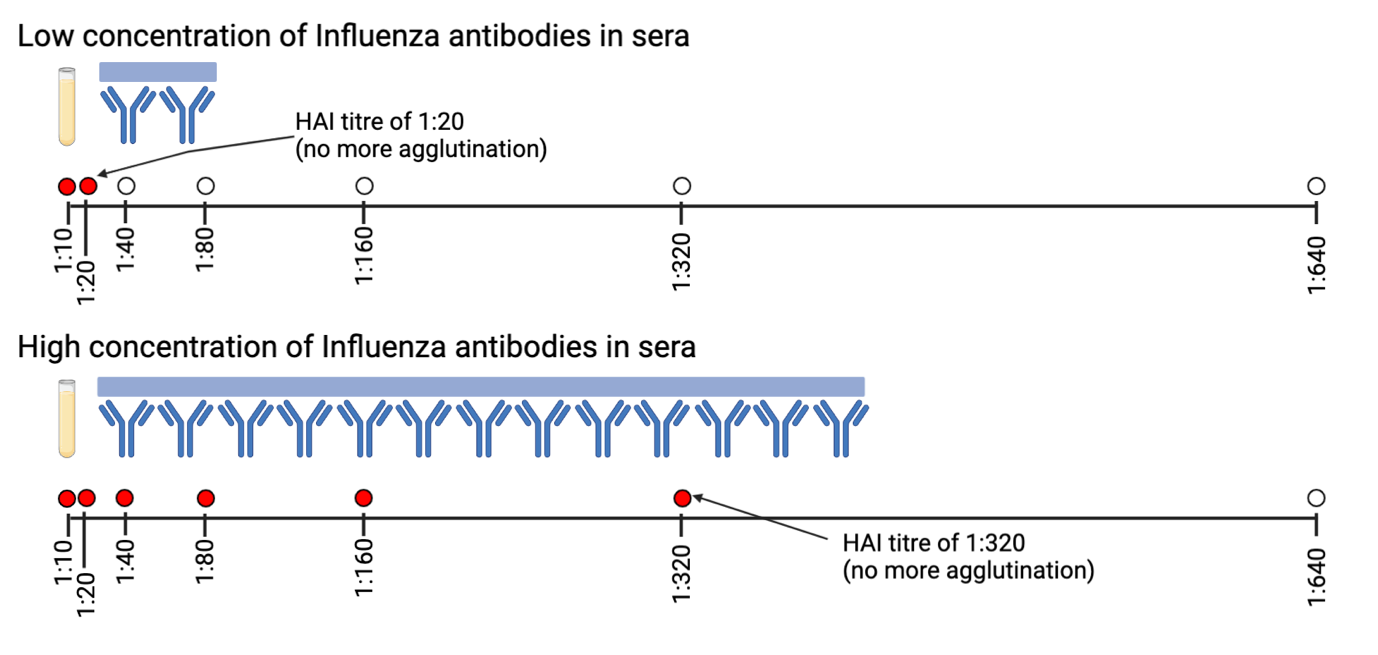

**Figure SM3**. Schematic showing the relationship between antibody titre in sera and HAI assay titre

| **Serum dilutions (**h_raw_) | <1:10 | 1:10 | 1:20 | 1:40 | 1:80 | 1:160 | 1:320 | 1:640 | 1:1280 | 1:2560 |
| --- | --- | --- | --- | --- | --- | --- | --- | --- | --- | --- |
| **Scaled Log_2_ dilutions (**x_i_) | 0 | 1 | 2 | 3 | 4 | 5 | 6 | 7 | 8 | 9 |

**Table SM1**. Equivalence between the serum dilutions and the scale log_2_ dilutions

1.2.2 Response variable

The response variable for our regression model is the fold-rise in HAI antibody titre (b_raw_) at time *t* post-vaccination (h_raw,t_) compared to the HAI antibody titre immediately before vaccination (h_raw,0_) (b_raw_ = h_raw,t_/ h_raw,0_). As we work on a log_2_ scale, fold-rise becomes the difference in log_2_ HAI antibody titre (b) at time *t* post-vaccination compared to the log_2_ HAI antibody titre immediately before vaccination. (b_i_ = x_i,t_ - x_i,0_).

1.2.3. Covariates

We consider the log_2_ HAI antibody titre immediately before vaccination as a covariate (x_i,0_). We expect, *a priori*, that log_2_ pre-vaccination HAI titre will be associated with log_2_ boosting post-vaccination. For pre-vaccination HAI titre to not have an associative effect, the fold-rise in antibodies would have to be the same independent for all pre-vaccination HAI titre. This is extremely unlikely as at high pre-vaccination titre levels, the data show attenuation in boosting on a log2 scale. This may be because there is an upper limit to the concentration of antibodies in the sera that can exist or be made in response to vaccination or because even if there is a substantial increase in antibody concentration, sera become so highly diluted in the HAI assay that this increase is not detectable.

In addition, we consider host factors, including age, gender, vaccine history and study site. Age could have a causal effect on antibody dynamics because of the effects of ageing on the immune system (immunosenescence).[1,2] Sex has also been shown to influence immune responses, potentially due to genetic mediators on the X chromosomes.[3] Finally, we consider vaccination history as a covariate influencing immune responses as it has been shown in various studies that highly vaccinated individuals have attenuated efficacy to infection compared to non-vaccinated individuals.[4] A table showing covariates in the model and their categoric or continuous values is shown in **Table SM2.**

| **Covariate** | **Data type** | **Range** |
| --- | --- | --- |
| Pre-vaccination titre | Categoric | {<1:10, 1:10, 1:20, 1:40, 1:80, 1:160, 1:320, 1:640, 1:1280, 1:2560} |
| Age groups | Categoric | {<30, 30–39, 40–49, 50–59, 60+} |
| Gender | Categoric | {Male, Female, Other} |
| Study site | Categoric | {Adelaide, Brisbane, Melbourne, Newcastle, Perth, Sydney} |
| Vaccine history (# of flu vaccines received in last 5 years) | Categoric | {0, 1, 2, 3, 4, 5} |

**Table SM2.** Summary of the covariates and their potential values.

1.3 Bayesian model outline

We developed a Bayesian model that estimates each participant's antibody response—as measured by HAI titre—at time *t* post-vaccination. The post-vaccine antibody kinetics model assumes an instantaneous boost immediately after vaccination, followed by a linear wane. Two latent parameters describe this model: the peak antibody boosting, *b*, (log_2_ scale) and the daily waning rate of antibody decline, *w*, (log_2_ scale). **Figure** **SM5** shows a directed acyclic diagram showing the relationship between the parameters in the Bayesian model, their deterministic and stochastic relationships, and the fitted data (HAI boost at time *t*).

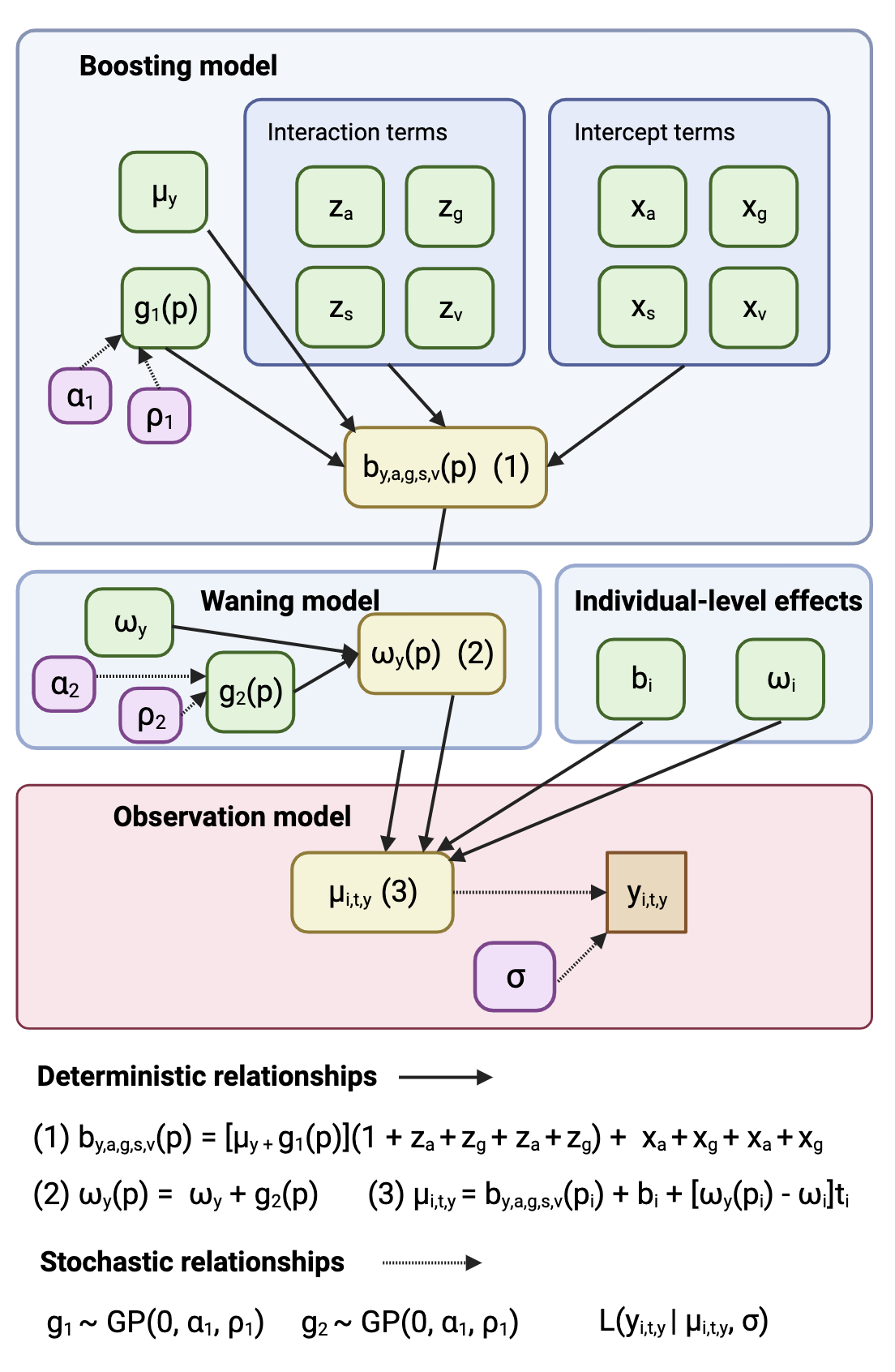

***Figure SM5.*** *Directed acyclic diagram showing the relationship between the parameters in the model, t their deterministic and stochastic relationships, and the fitted data (HAI boost at time t). Parameters in purple boxes are fitted hyperparameters, parameters in the green box show a partial pooling structure and are stochastically determined through their parent nodes (not all are shown; see* ***Table S2****), parameters in yellow boxes are deterministically determined from their parent nodes, and the orange box refers to the data. Subscripts y, a, g, s, and v, refer to years of study, age group, gender, study site, and vaccination history, respectively, and are all categoric variables. Subscript, i, refers to an individual. The parameter p is the pre-vaccination titre, and the parameter t refers to the time post-vaccination.* ***GP*** *refers to a Gaussian process prior and f is the likelihood function.*

The likelihood is chosen to convert continuous estimates of the antibody concentration from the regressions model ($\mu_{i, t,y}$) into discrete observed titre boost $y_{i, t,y}$:

$$L\left( y_{i,t,y} | \mu_{i,t,y}, \sigma\right)=\int_{t_{i,t,t}-1}^{y_{i,t,y}+1} f(x,|\mu_{i,t,y}, \sigma)dx$$

Where $f(x,|\mu_{i,t,y}, \sigma)$ is the probability density function of the normal distribution with mean $\mu_{i,t,y}$ and standard deviation $\sigma$.

A full description of each of the variables shown in **Figure S5**, along with their prior distributions, is given in **Table SM3**.

| **Symbol** | **Description** | **Prior** |
| --- | --- | --- |
| *Data* | | |
| $y_{i,t,y}$ | HAI titre value at time *t* for individual i during year *y*. | — |
| $p_{i}$ | Pre-vaccination HAI titre value for individual *i* | — |
| $a_{i}$ | Age group for individual *i* | — |
| $g_{i}$ | Gender of individual *i* | — |
| $s_{i}$ | Study site for individual *i* | — |
| $v_{i}$ | Vaccination history for individual *i* |  |
| *Antibody boosting model* | | |
| $\mu_{y}$ | Mean peak antibody boosting in study year *y* (log_2_) | $N(3, 2)$ |
| $\alpha_{1}$ | Scale parameter in Gaussian process prior model for boosting | $N^{+}(0, 1)$ |
| $\rho_{1}$ | Length parameter in Gaussian process prior model for boosting | $Inv\_Gamma(5, 5)$ |
| $x_{a}$ | Intercepts for categorical age groups data, pooled effects | $N(0, \sigma_{a1})$  $\sigma_{a1}\sim N^{+}(0, 1)$ |
| $x_{g}$ | Intercepts for categorical gender data, pooled effects | $N(0, \sigma_{g1})$  $\sigma_{g1}\sim N^{+}(0, 1)$ |
| $x_{s}$ | Intercepts for categorical study-site data, pooled effects | $N(0, \sigma_{s1})$  $\sigma_{s1}\sim N^{+}(0, 1)$ |
| $x_{v}$ | Intercepts for categorical vaccine history data, pooled effects | $N(0, \sigma_{v1})$  $\sigma_{v1}\sim N^{+}(0, 1)$ |
| $z_{a}$ | Interaction term for categorical age groups data, pooled effects | $N(0, \sigma_{a2})$  $\sigma_{a2}\sim N^{+}(0,0.1)$ |
| $z_{g}$ | Interaction term for categorical gender data, pooled effects | $N(0, \sigma_{g2})$  $\sigma_{g2}\sim N^{+}(0, 0.1)$ |
| $z_{s}$ | Interaction term for categorical study-site data, pooled effects | $N(0, \sigma_{s2})$  $\sigma_{s2}\sim N^{+}(0, 0.1)$ |
| $z_{v}$ | Interaction term for categorical vaccine history data, pooled effects | $N(0, \sigma_{v2})$  $\sigma_{v2}\sim N^{+}(0, 0.1)$ |
| *Antibody waning model* | | |
| $\omega_{y}$ | Mean antibody waning per day in study year *c* (log_2_) | $N(-0.003, 0.002)$ |
| $\alpha_{2}$ | Scale parameter in Gaussian process prior model for waning | $N^{+}(0, 0.002)$ |
| $\rho_{3}$ | Length parameter in Gaussian process prior model for waning | $Inv\_Gamma(5, 5)$ |
| *Individual-level effects* | | |
| $b_{i}$ | Individual-level effect (deviations from the population model) for peak boosting | $N(0, \sigma_{b})$  $\sigma_{a2}\sim N(0,1)$ |
| $w_{i}$ | Individual-level effect (deviations from the population model) for waning per day | $N(0, \sigma_{w})$  $\sigma_{w}\sim N(0,0.001)$ |
| *Likelihood function* | | |
| $y_{i,t}$ | Estimated antibody concentration boost for individual *i* at time *t* | See Above. |
| $\sigma$ | Standard deviation of normal distribution in likelihood | *Exponential(1)* |

**Table SM3.** Summary of parameters and prior distributions in the model.

1.3.1 Justification for Gaussian Process Prior

We assume that pre-vaccination HAI titre has a significant impact on boosting. These values are discrete but ordered. Therefore, we assume they follow a latent variable Gaussian process prior (GP). Due to the complexity of immunological mechanisms that relate to pre-vaccination HAI titre and boosting, fitting using a non-parametric form such as Gaussian process prior allows for a flexible non-linear relationship to be established (see **Figure SM5**). Further, as the extremes of the HAI titre values have a low number of samples, a Gaussian process prior will inform these titre values with low power from similar values, giving a more sensible estimate than alternative models, such as a random-effect model, might give (**Figure SM6**).

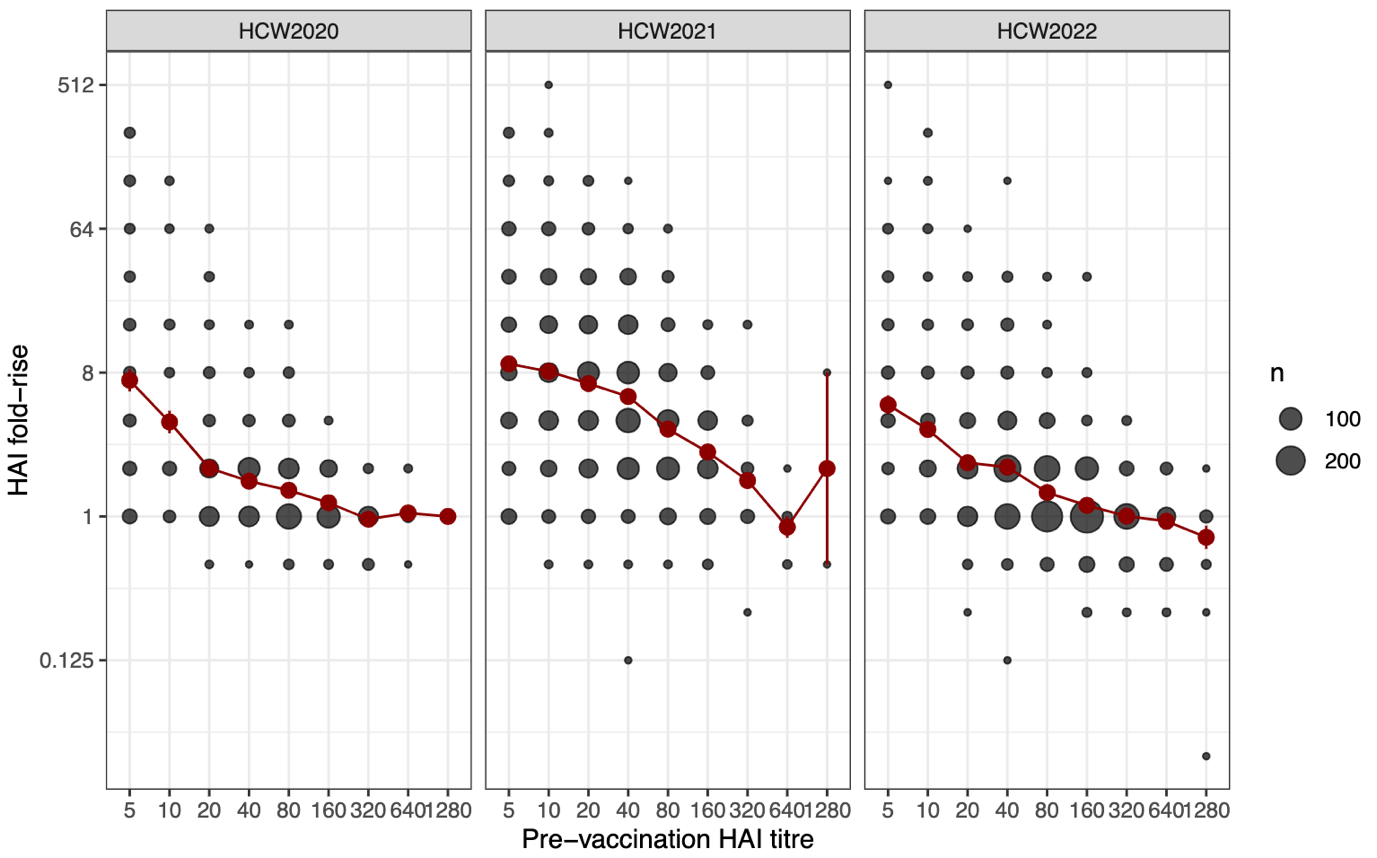

**Figure SM5.** HAI boosting and Pre-vaccination HAI titre value for each study year.

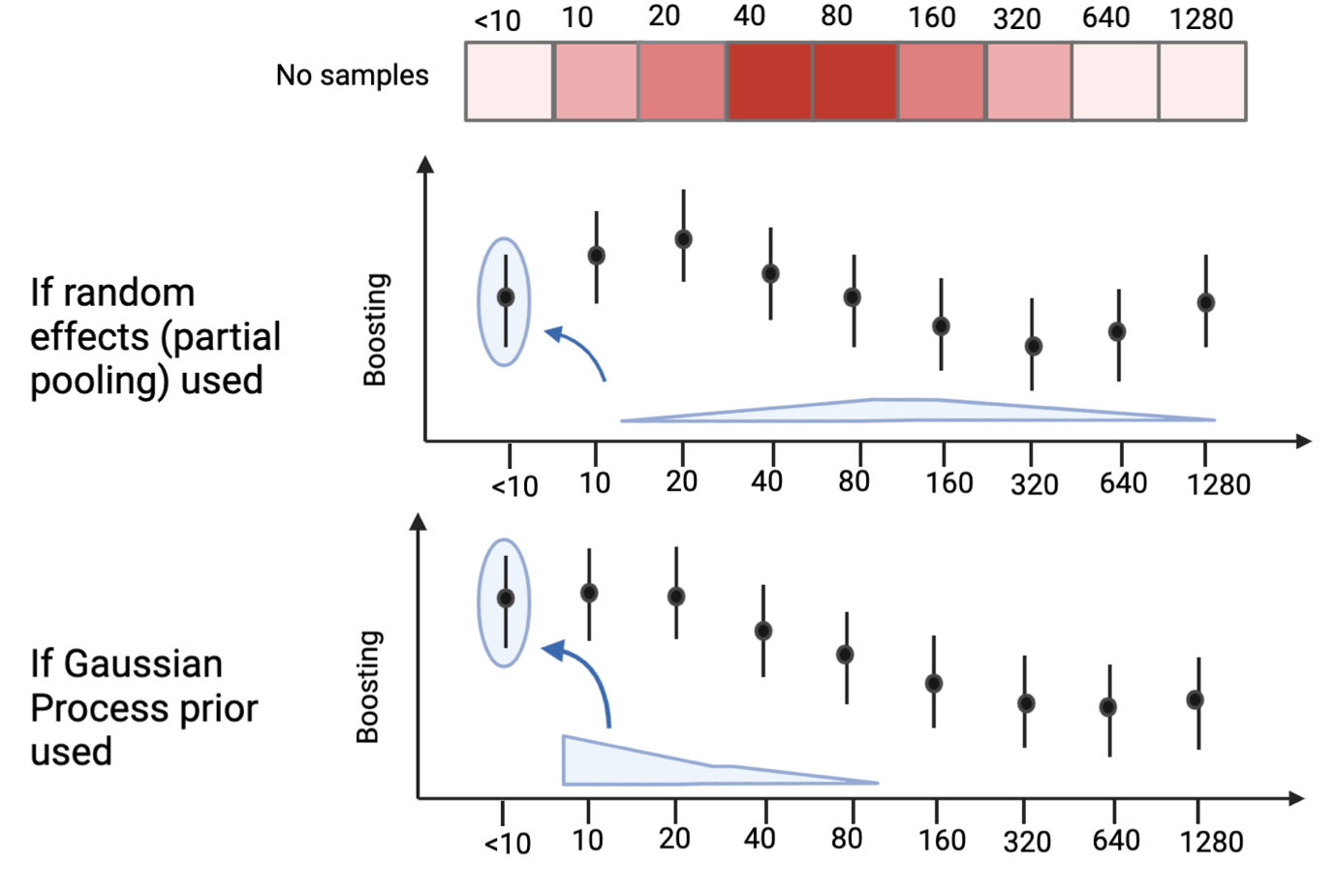

**Figure SM6.** Schematic showing the justification for the Gaussian process prior in comparison to a partial pooling model.

1.4 Implementation

Our Bayesian model highlighted in Section 1.3 was coded and fitting using Hamiltonian Monte Carlo (HMC) using stan. We ran the HMC with 4 chains with 2,000 steps each, 1,000 of which were burn-in.

Supplementary Figures to “A Bayesian model for the in-host antibody kinetics driven by seasonal influenza vaccination”

2.1 Posterior predictive distributions vs data

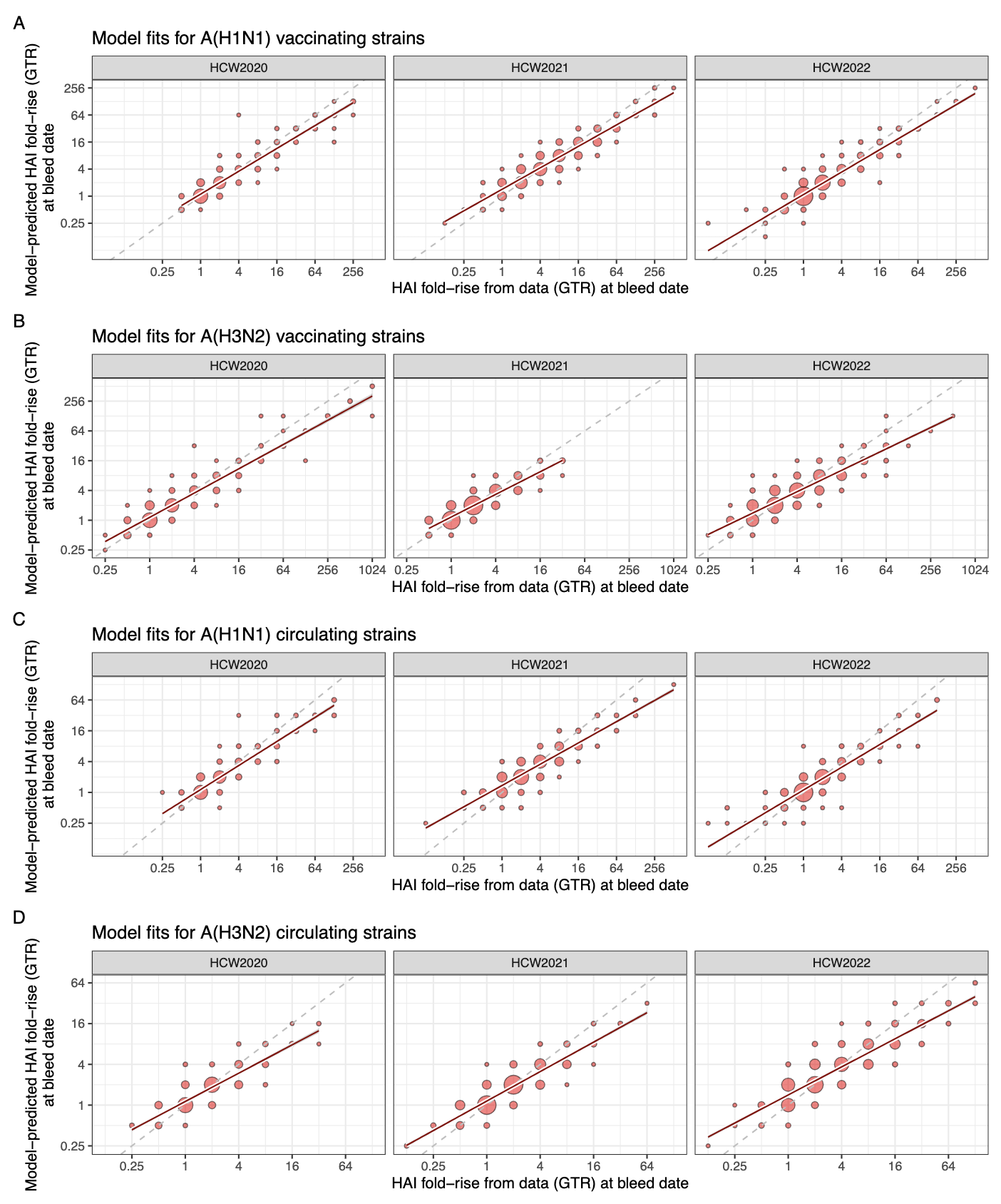

Figure S1. **Summary of the Bayesian model-fits to the serological data.** The size of the point marker is scaled to the number of samples, the dashed line marks where the models are equal, and the fitted line is a simple linear regression. (A) shows the fits for the A(H1N1) vaccinating viruses (B) shows the fits for the A(H3N2) vaccinating viruses, (C) shows the fits for the A(H1N1) viruses and (D) shows the fits for A(H3N2) circulating viruses; all four are stratified by study year.

2.1 Vaccine-induced kinetics of A(H1N1) vaccinating strains

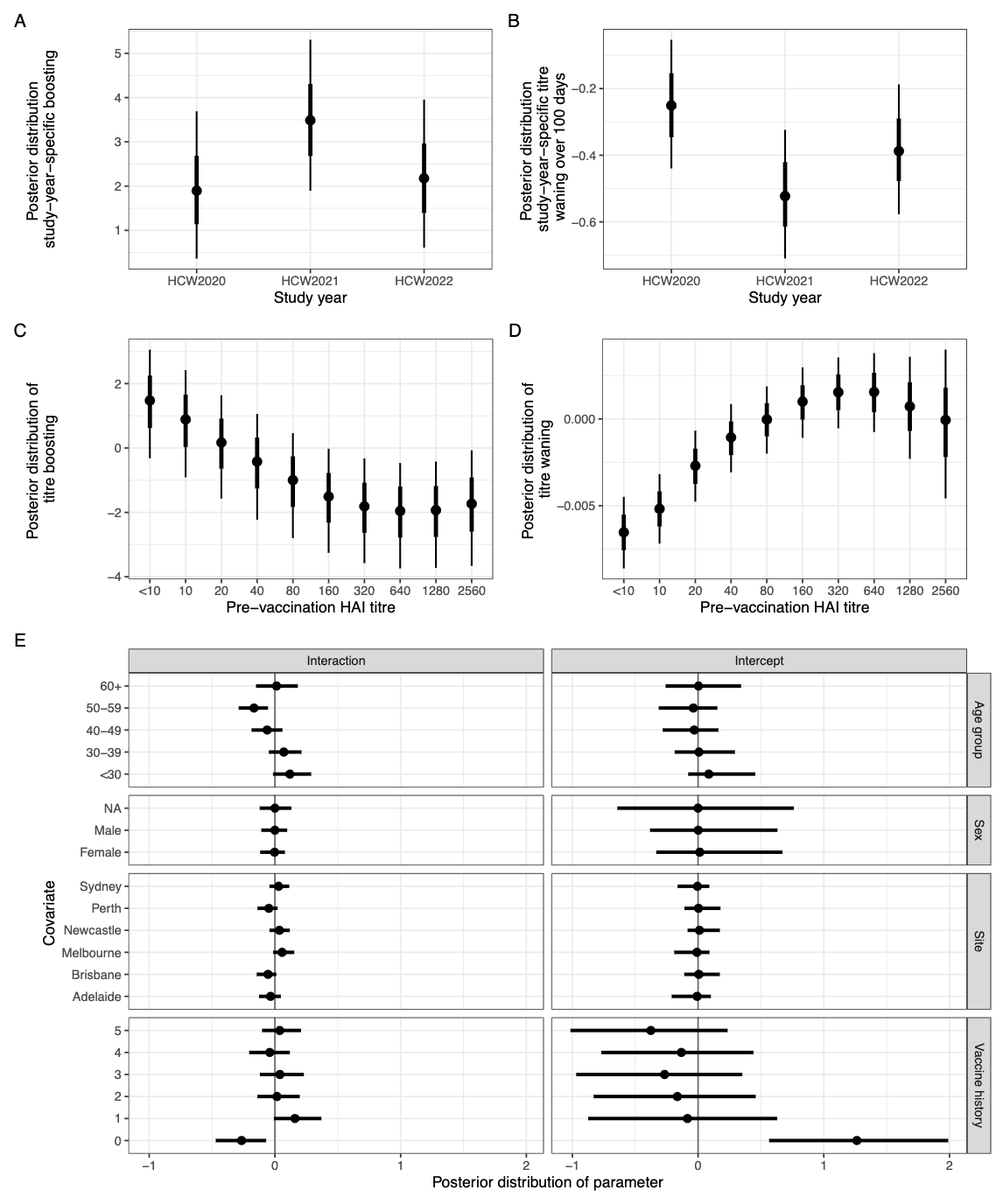

**Figure S2.** Posterior distribution of the effect size for the A(H1N1) vaccinating strains

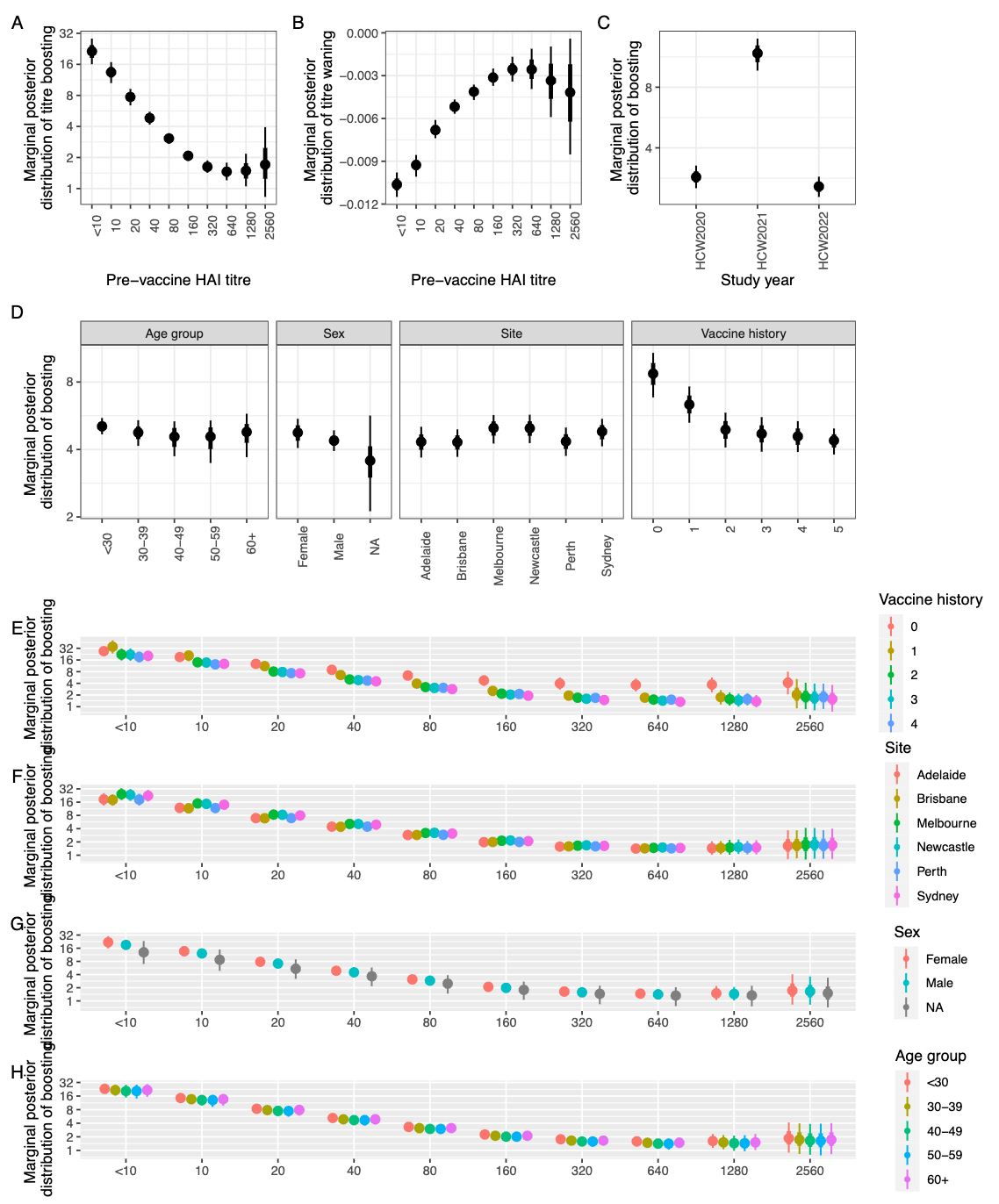

**Figure S3.** The marginal posterior distribution for the A(H1N1) vaccinating strains

2.2 Vaccine-induced kinetics of A(3N2) vaccinating strains

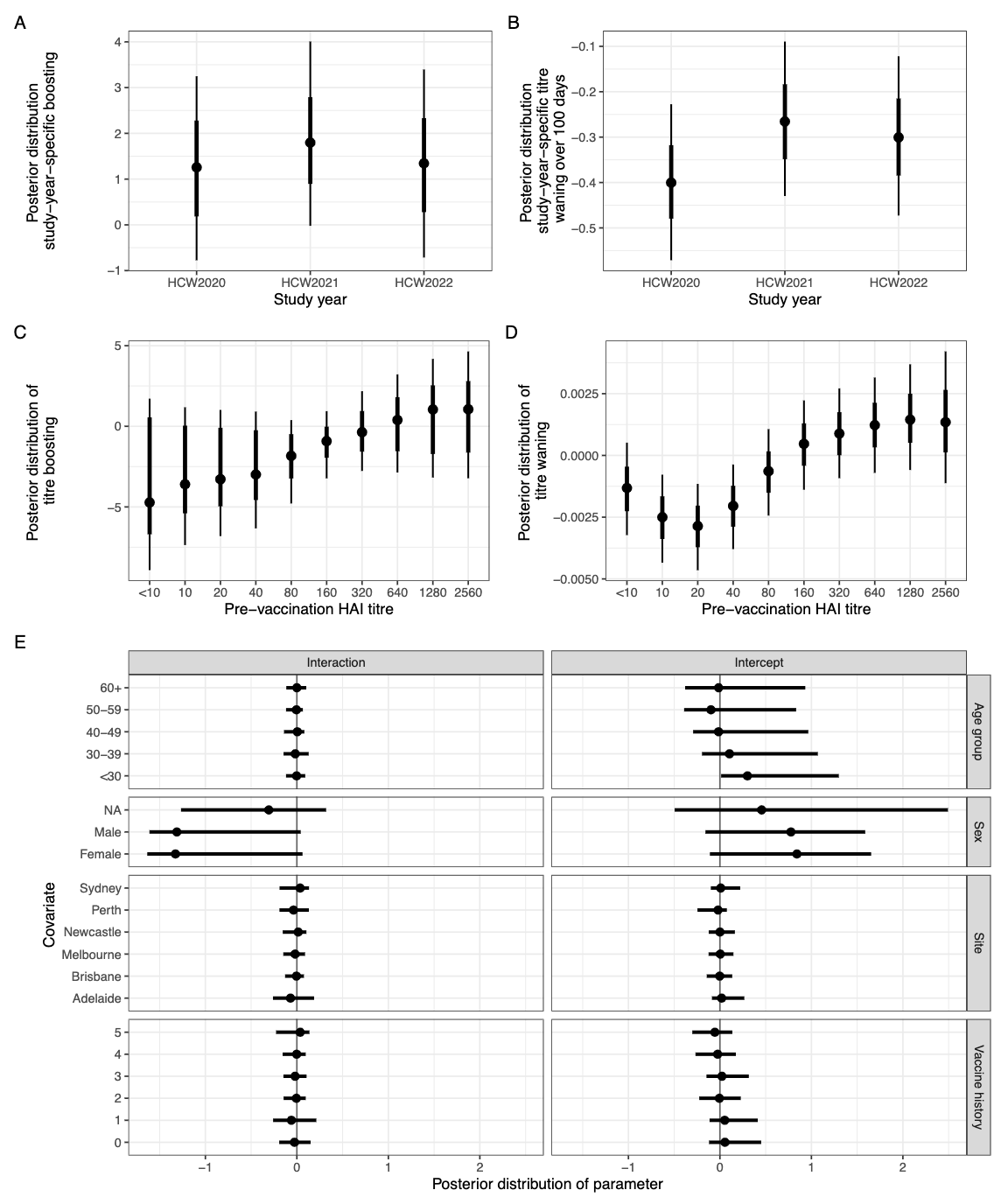

**Figure S4.** Posterior distribution of the effect size for the A(H3N2) vaccinating strains

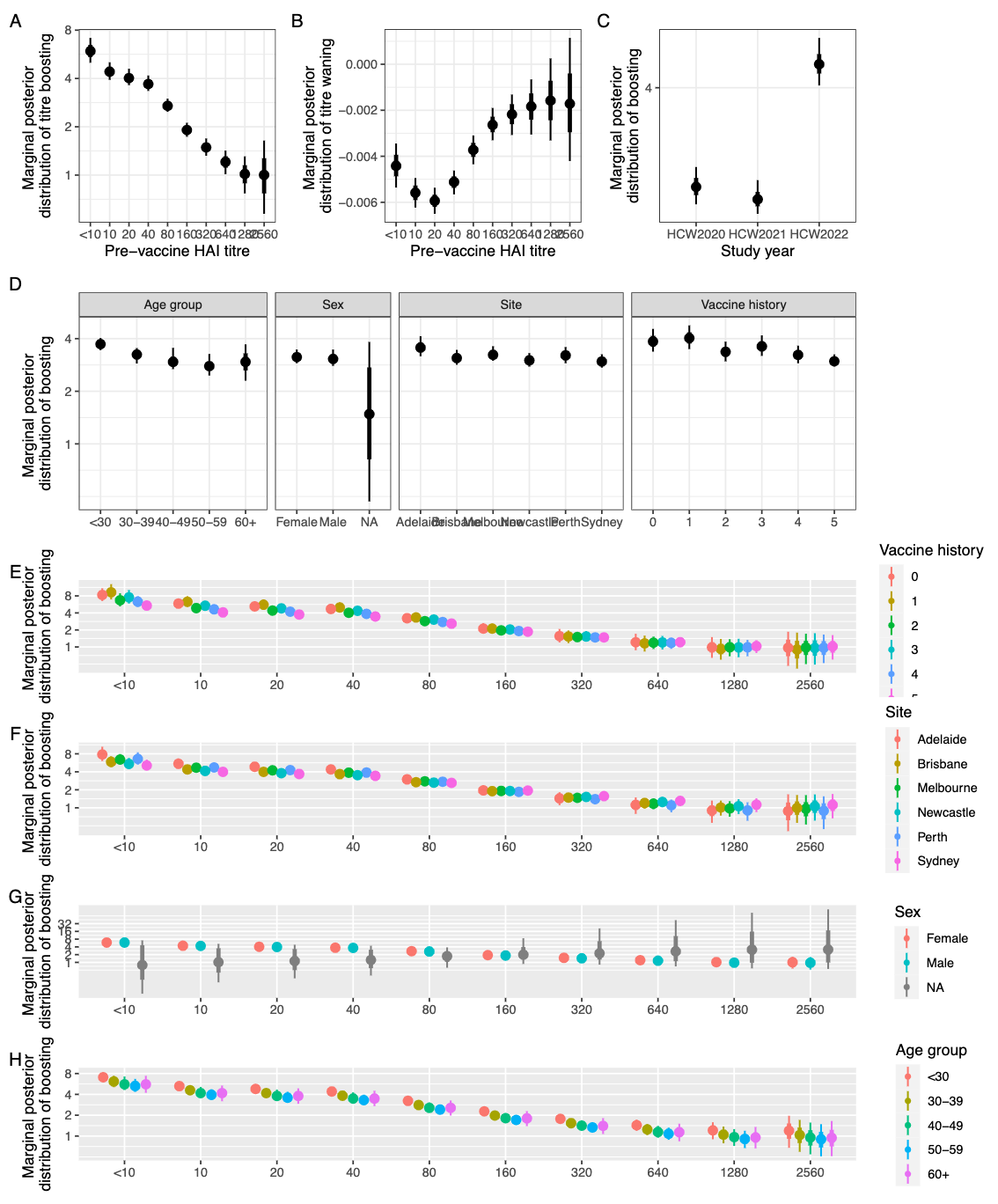

**Figure S5.** The marginal posterior distribution for the A(H3N2) vaccinating strains

2.3. Vaccine-induced kinetics of A(H1N1) circulating strains

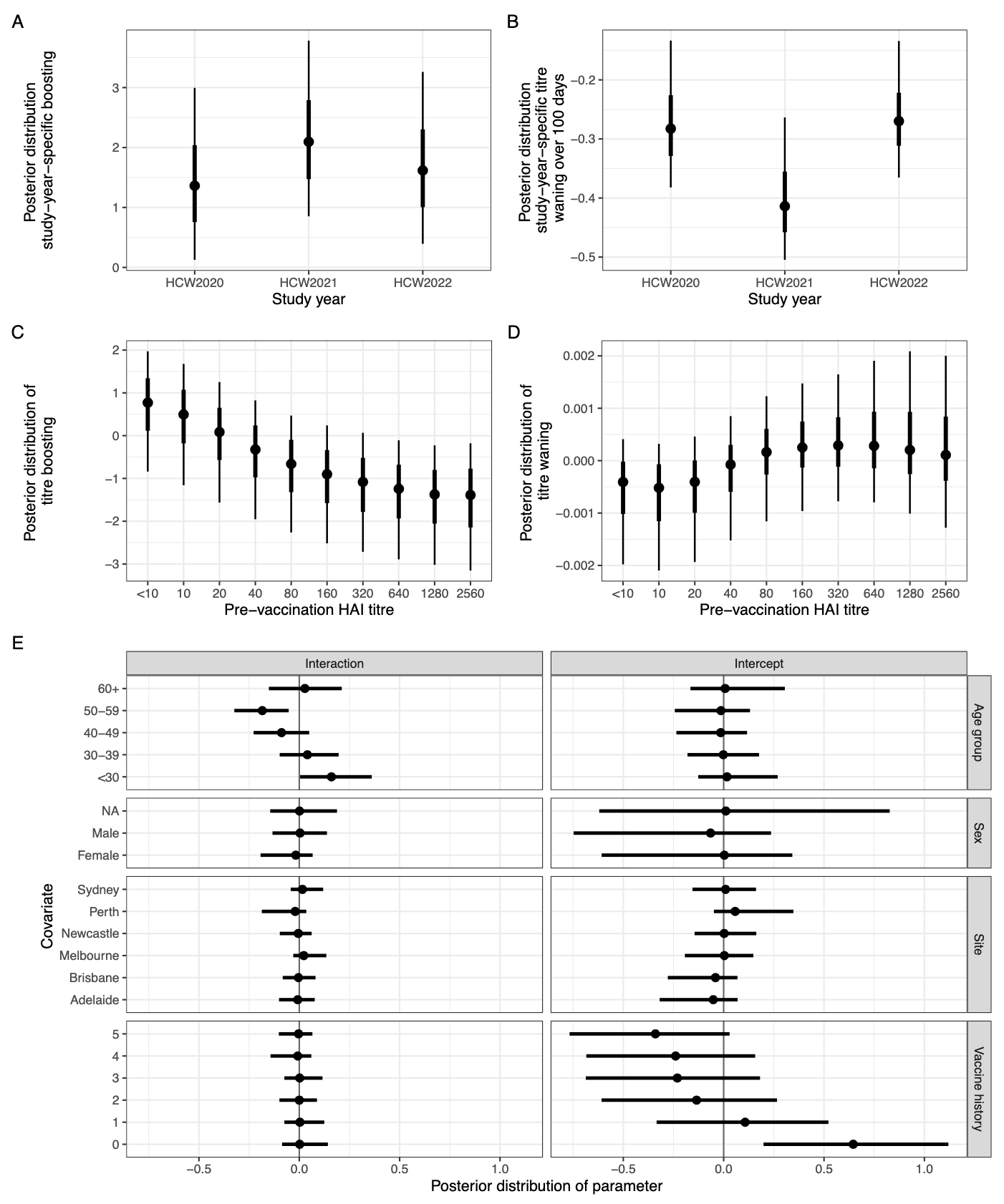

**Figure S6.** Posterior distribution of the effect size for the A(H1N1) circulating strains

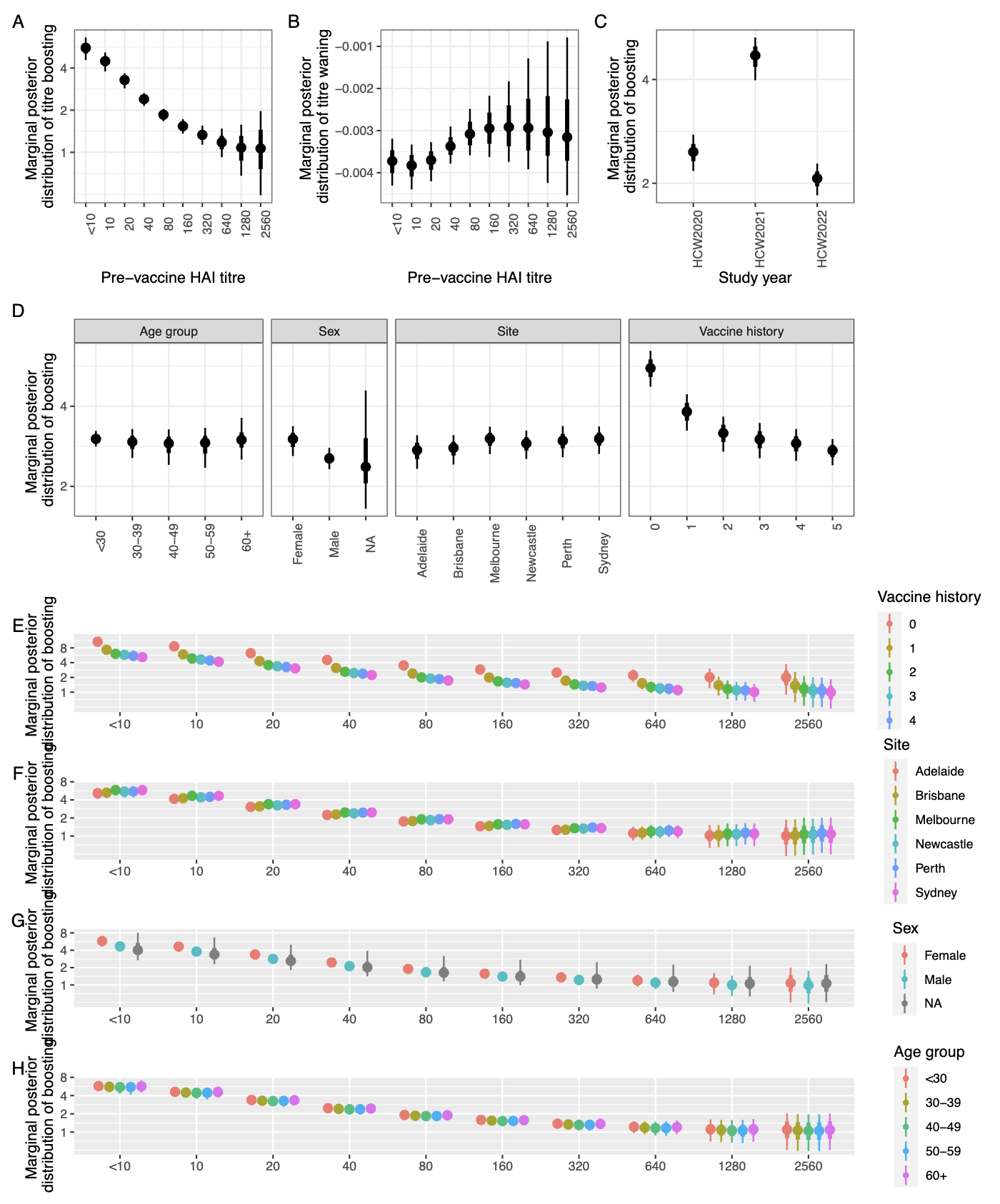

**Figure S7.** The marginal posterior distribution for the A(H1N1) circulating strains

2.4. Vaccine-induced kinetics of A(H3N2) circulating strains

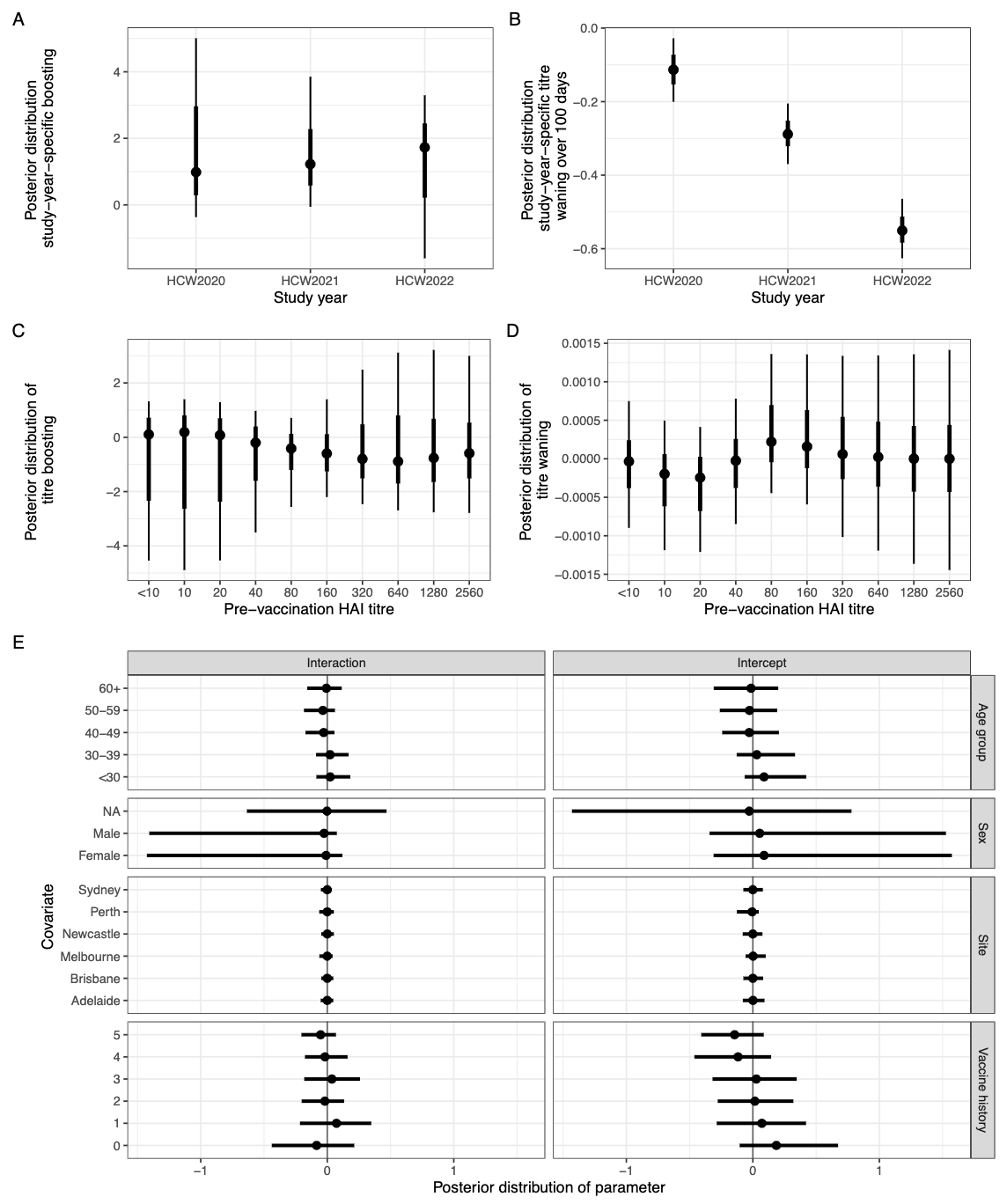

**Figure S8.** Posterior distribution of the effect size for the A(H3N2) circulating strains

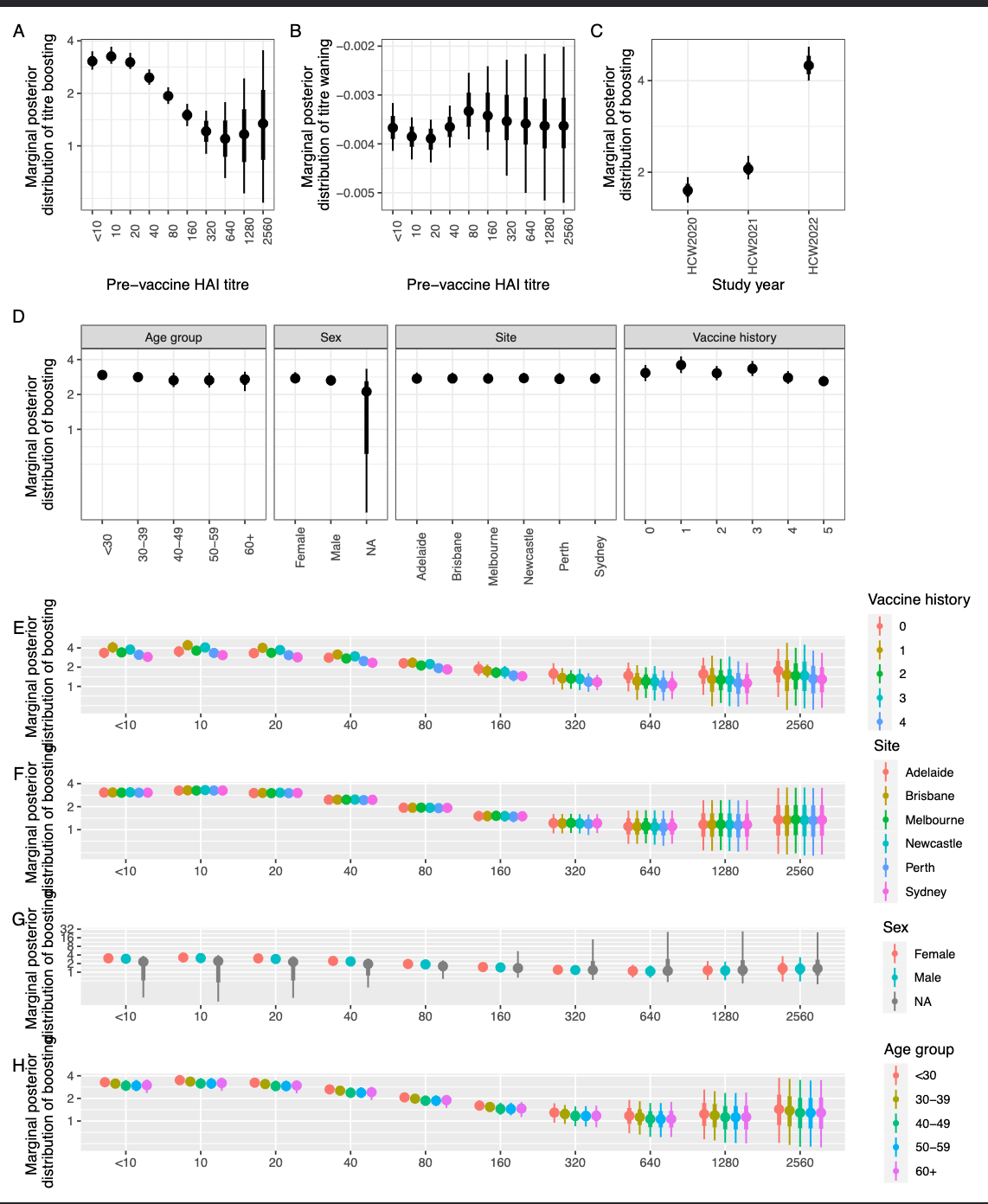

**Figure S9.** The marginal posterior distribution for the A(H3N2) circulating strains

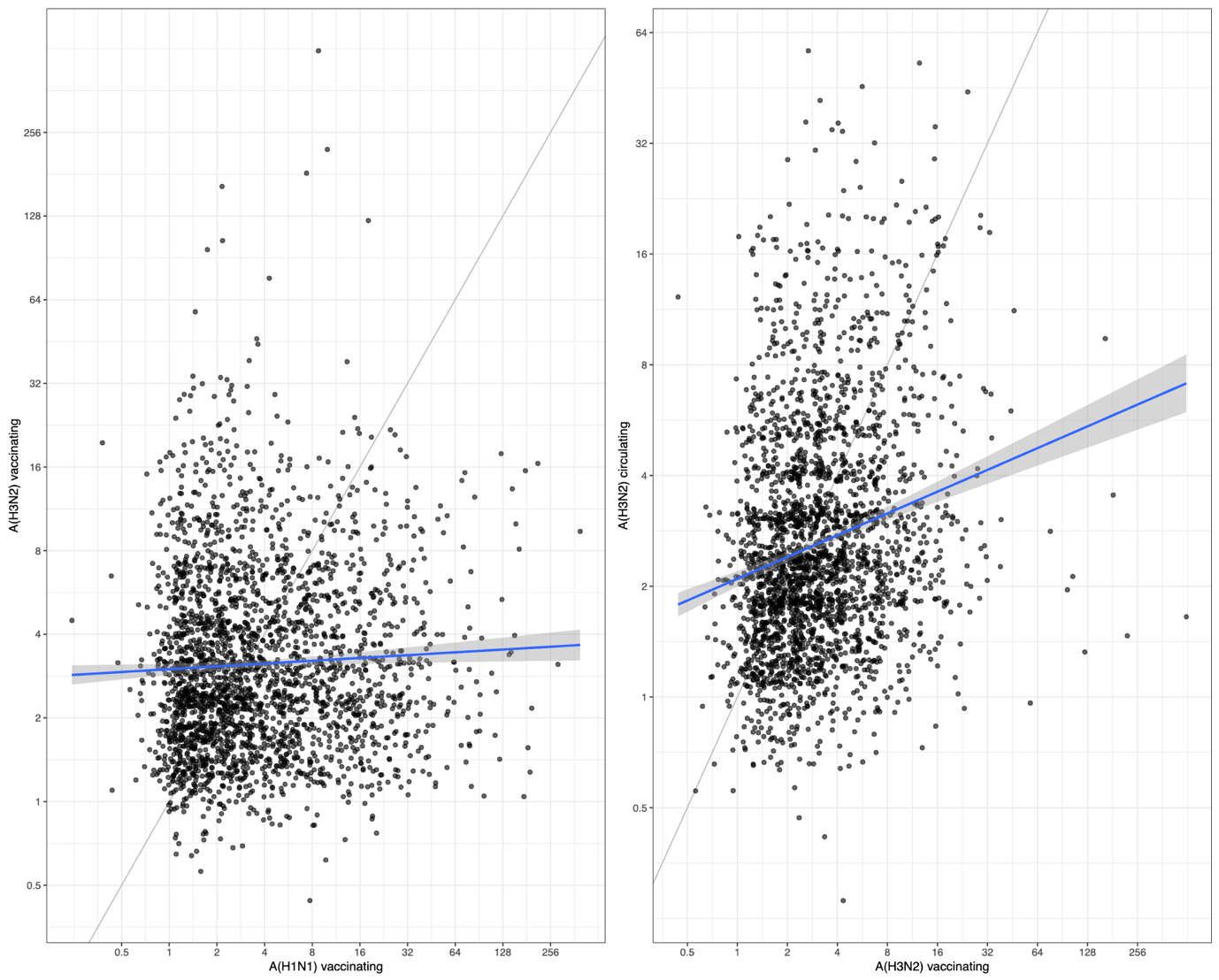

**Figure S10.** Plot of the individual-level model-estimated outcome variability between strain subtypes

Supplementary Tables to “A Bayesian model for the in-host antibody kinetics driven by seasonal influenza vaccination”

| *Season* | *Cohort* | *H3 vaccine antigen* | *H1 vaccine antigen* |
| --- | --- | --- | --- |
| *2019* | *NA* | *A/Switzerland/8060/2017* | *A/Brisbane/02/2018* |
| *2019/20* | *NA* | *A/Kansas/14/2017* | *A/Brisbane/02/2018* |
| *2020* | *Australia* | *A/South Australia/34/2019* | *A/Brisbane/02/2018* |
| *2021* | *Australia* | *A/Hong Kong/2671/2019* | *A/Victoria/2570/2019* |
| *2022* | *Australia* | *A/Darwin/9/2021* | *A/Victoria/2570/2019* |

*Supplementary Table 1. Influenza A vaccine antigens received during the study period.*
